# Selection of artemisinin partial resistance Kelch13 mutations in Uganda in 2016-22 was at a rate comparable to that seen previously in South-East Asia

**DOI:** 10.1101/2024.02.03.24302209

**Authors:** Cecile P. G. Meier-Scherling, Oliver J Watson, Victor Asua, Isaac Ghinai, Thomas Katairo, Shreeya Garg, Melissa Conrad, Philip J. Rosenthal, Lucy C Okell, Jeffrey A. Bailey

## Abstract

**Background:** Artemisinin partial resistance, mediated by mutations in the *Plasmodium falciparum* Kelch13 protein (K13), rapidly spread in South-East Asia (SEA), undermining antimalarial efficacies of artemisinin-based combination therapies (ACT). Validated K13 mutations have recently arisen in Africa, but rates of increase are not well characterized.

**Methods:** We investigated K13 mutation prevalence at 16 sites in Uganda (2016-2022, 6586 samples), and five sites in SEA (2003-2018, 5465 samples) by calculating selection coefficients using Bayesian mixed-effect linear models. We then tested whether SEA K13 mutation prevalence could have been forecast accurately using up to the first five years of available data and forecast future K13 mutation prevalence in Uganda.

**Findings:** The selection coefficient for the prevalence of relevant K13 mutations (441L, 469F/Y, 561H, 675V) was estimated at s=0·383 (95% CrI: 0·247 - 0·528) per year, a 38% relative prevalence increase. Selection coefficients across Uganda were s=0·968 (0·463 - 1·569) for 441L, s=0·153 (- 0·445 - 0·727) for 469F, s=0·222 (−0·011 - 0·398) for 469Y, and s=0·152 (−0·023 - 0·312) for 675V. In SEA, the selection coefficient was s=−0·005 (−0·852 - 0·814) for 539T, s=0·574 (−0·092 - 1·201) for 580Y, and s=0·308 (0·089 - 0·536) for all validated K13 mutations. Forecast prevalences for Uganda assuming constant selection neared fixation (>95% prevalence) within a decade (2028-2033) for combined K13 mutations.

**Interpretation:** The selection of K13 mutations in Uganda was at a comparable rate to that observed in SEA, suggesting K13 mutations may continue to increase quickly in Uganda.

**Funding:** NIH R01AI156267, R01AI075045, and R01AI089674.

## Research in Context

### Evidence before this study

Artemisinin partial resistance, mediated by K13 propeller mutations, has been confirmed in multiple locations in Eastern Africa. However, longitudinal data is limited; consequently, it still has to be quantified how quickly these mutations spread over time and how they will evolve.

### Added value of this study

To quantify the current spread of mutations at multiple sites in Uganda, we fitted Bayesian mixed-effect linear models to estimate the selection coefficients of K13 mutations associated with artemisinin partial resistance (ART-R). Selection coefficients quantify the change in the relative prevalence of a relevant genotype. Comparing those estimates to the early spread of resistance in South-East Asia (SEA), we found that rates of selection in Uganda were comparable to those during the early spread of resistance in SEA, where ART-R is now widespread. Further, we forecast the prevalence of ART-R in SEA under the assumption of constant selection, using data from the first five years of the emergence of resistance to quantify the accuracy of forecasting resistance prevalence in this way. Compared to the observed data in SEA, the forecast prevalence underestimated the true prevalence by a weighted mean error of 16%. We also used this method with available data in Uganda, predicting near fixation (>95% prevalence) of ART-R mutations within ten years.

### Implications of all the available evidence

Our modeling suggests that markers of ART-R are increasing in Uganda at rates comparable to those seen previously during the early stages of ART-R emergence in SEA.

## Background

The World Health Organization (WHO) has recommended artemisinin-based combination therapies (ACTs) for treating uncomplicated malaria since 2001. ACTs comprise a fast-acting artemisinin derivative for rapid parasite reduction and a longer-acting partner drug that clears remaining parasites.^1–5^ In 2008, artemisinin partial resistance (ART-R) was first reported in Cambodia and later determined to have emerged around 2001.^6,7^ ART-R is caused by mutations in the *Plasmodium falciparum* Kelch13 protein (K13) propeller domain.^7^ *In vitro* and *in vivo* evidence suggest that parasites carrying K13 mutations are less fit than wild-type parasites without drug pressure, indicating likely selective advantages under current treatment and transmission dynamics.^8,9^ In SEA, K13 mutation spread was quickly accompanied by resistance to ACT partner drugs, notably mefloquine and piperaquine. Thus, there is concern that if resistance-mediating K13 mutations spread rapidly in Africa, partner drug resistance and clinical treatment failures will soon follow.

Recent studies have shown that ART-R has emerged independently in multiple countries in eastern Africa. De novo emergence of the K13 561H mutation was first identified in Rwanda in 2014.^10^ The 469Y and 675V mutations were identified in isolates collected in Uganda beginning in 2016 and proven to enhance parasite survival *in vitro*.^11,12^ Shortly after, 469Y and 675V mutations were associated with partial resistance clinically and *in vitro*.^12–15^ The 622I mutation was reported in Eritrea in samples from 2016-2019 and Ethiopia as early as 2014.^16–18^ The study in Eritrea also found an increased risk of parasitemia three days after initiation of treatment amongst patients with 622I mutated parasites.^16^ To date, the K13 propeller domain mutations that have shown appreciable site prevalences (>10%) and spread in Africa are all validated or candidate mutations based on the WHO schema.^19^

In Uganda, data from 10-16 sites have been collected annually since 2016, assessing drug-resistance mutations in isolates causing uncomplicated malaria.^20^ Interestingly, the emergence of ART-R in Uganda differed from its emergence in SEA in some respects. Resistance first emerged in northern Uganda with historically high levels of malaria transmission, contrasting the emergence in low transmission regions of SEA.^20^ However, heterogeneity in malaria control measures, including indoor residual spraying of insecticides, may explain these patterns.^20^ Given Africa’s overall higher and greater transmission range, the ART-R in Africa may differ significantly from those observed in SEA.^20^ To facilitate cross-study comparisons of resistance dynamics and to predict the future spread of resistance, selection coefficients (s) are estimated, which represent the percent change in the relative prevalence of the mutant genotype per unit time, quantifying if a genetic variant provides an advantage (s>0) or disadvantage (s<0).^1,21^ Understanding these dynamics can aid in designing treatment policies to slow the spread of resistance.^22^ Here, we fit Bayesian mixed-effect linear models to estimate selection coefficients in 13 Ugandan districts with K13 mutations observed from 2016-2022 and compare these estimates with early SEA ART-R emergence in five SEAn sites in which K13 mutations were observed from 2003-2018.^20,23^

## Methods

### Study Sites for Uganda

The longitudinal K13 mutation prevalence data used for this study were generated by targeted sequencing of DNA from blood samples from individuals presenting with uncomplicated malaria (50 isolates/site for ten sites, 2016-2019; 100 isolates/site for 16 sites, 2020-2022) (Figure S1).^20^ Five validated/candidate markers of ART-R were observed: 441L, 469F/Y, 561H, and 675V.

### Study Sites for SEA

To compare estimated selection coefficients in Uganda and SEA, where ACT-R is widely spread, we estimated selection coefficients using the MalariaGen Pf7 dataset, with 5465 SEA samples.^23^ However, due to piperaquine partner drug resistance in later years in SEA, we focused on the initial increase in ART-R to allow for more suitable comparisons with Uganda.^24^ At each site, we only included the first five years of non-zero mutation prevalence when at least three of the five years had sampling. In addition, for inclusion, the first year of non-zero prevalence had to occur no later than 2010, avoiding later years when piperaquine partner drug resistance was most likely spreading in concert. The resulting SEA data covered three countries: Cambodia (three districts), Thailand (one district), and Vietnam (one district).

### Comparison of selection in Uganda and SEA

To estimate selection coefficients, we fit a Bayesian linear model to the logit-transformed mutation prevalence against time with a random slope and intercept for each site. We analyzed sites and mutations with non-zero mutation prevalence observed in at least three years. The mixed effects model allows the selection coefficient to vary between sites while leveraging information across sites. The model was weighted by the inverse variance of the logit-prevalence from each observation.^1^ We used the *rstanarm* package in R (version 4.2.1) to fit the models, using the default prior distributions for the fixed and random effects.^25^

In Uganda, K13 561H was observed in too few districts and years to meet the model’s inclusion criteria and thus was included in analyses based on all observed K13 mutations but not in individual mutation analyses. Using this model, we estimated selection coefficients across Uganda and all individual sites for individual K13 mutations (441L, 469F/Y, 675V) and all observed K13 mutations combined (441L, 469F/Y, 561H, 675V).

In SEA, we fit the model to estimate selection coefficients for two of the most prevalent SEA K13 mutations, 580Y and 539T, and the overall prevalence of all WHO-validated K13 mutations (446I, 458Y, 469Y, 476I, 493H, 539T, 543T, 553L, 561H, 574L, 580Y, 622I, 675V).

### Forecasting of selection in SEA and Uganda

We tested whether future trends in K13 prevalence can be predicted using data from the early stages of resistance emergence. To achieve this, we fit the same statistical model described above to the first three, four, or five consecutive years of non-zero mutation prevalence data in SEA. We used these models to forecast mutation prevalence in subsequent years, assuming constant selection over time. To focus on initial spread within sites in SEA, we only forecast prevalence in districts with <50% resistance prevalence in the first sampled year.

Using the fitted model estimates, we forecast mutation prevalence until 2023 for the most prevalent mutation (580Y) and 13 mutations combined (446I, 458Y, 469Y, 476I, 493H, 539T, 543T, 553L, 561H, 574L, 580Y, 622I, 675V) in SEA. Due to a lack of consecutive samples, 539T was not included in the individual mutation forecasting. To obtain the 95% posterior credible interval (CrI) for the selection coefficient, we sampled 100 draws from the posterior distribution and reported the 95% interpercentile distribution.^26^ We evaluated the forecast prevalence against observed data not used in the model (i.e., in forecast years) by calculating the correlation, mean absolute error, and bias estimates.

Similar to the forecasting in SEA, we sampled 100 draws from the posterior distribution of fitted statistical models to forecast K13 mutation prevalence for each evaluable Ugandan mutation (441L, 469F/Y, 675V) and all five observed K13 mutations overall (441L, 469F/Y, 561H, 675V) in Uganda, assuming constant selection over time.

### Literature Review

To compare against selection coefficients for other mutations, we searched PubMed up to 29 November 2023 using the search terms: (“selection coefficient” OR “selective sweep” OR “spread of antimalarial drug resistance”[tiab:∼0] OR “changes in malaria parasite drug resistance”[tiab:∼0] OR “impact of antimalarial drug resistance”[tiab:∼0]) AND (malaria[MESH] OR malaria[Text word]) AND (antimalarial drug resistance). Identified publications were screened for reported selection coefficients from clinical samples. We standardized all identified selection coefficients by converting to per year rather than per parasite generation.

## Results

### K13 selection in Uganda is comparable to early selection in SEA

We obtained annual K13 mutation surveillance data from 2016-2022, totaling 6586 isolates from patients presenting with uncomplicated malaria at sites across Uganda (Figure S1, Table S1).^20^ These included five different K13 mutations that are validated/candidate markers of ART-R (441L, 469F/Y, 561H, 675V). Overall K13 mutation prevalence increased from 4·5% (15/330) in 2016 to 25·5% (256/1003) in 2022 (Figure 1A).^20^ K13 469Y and 675V were mostly observed in northern Uganda, while 441L, 469F, and 561H were most prevalent in western and south-western Uganda (Figure 1B, Figure S2).

**Figure 1.**
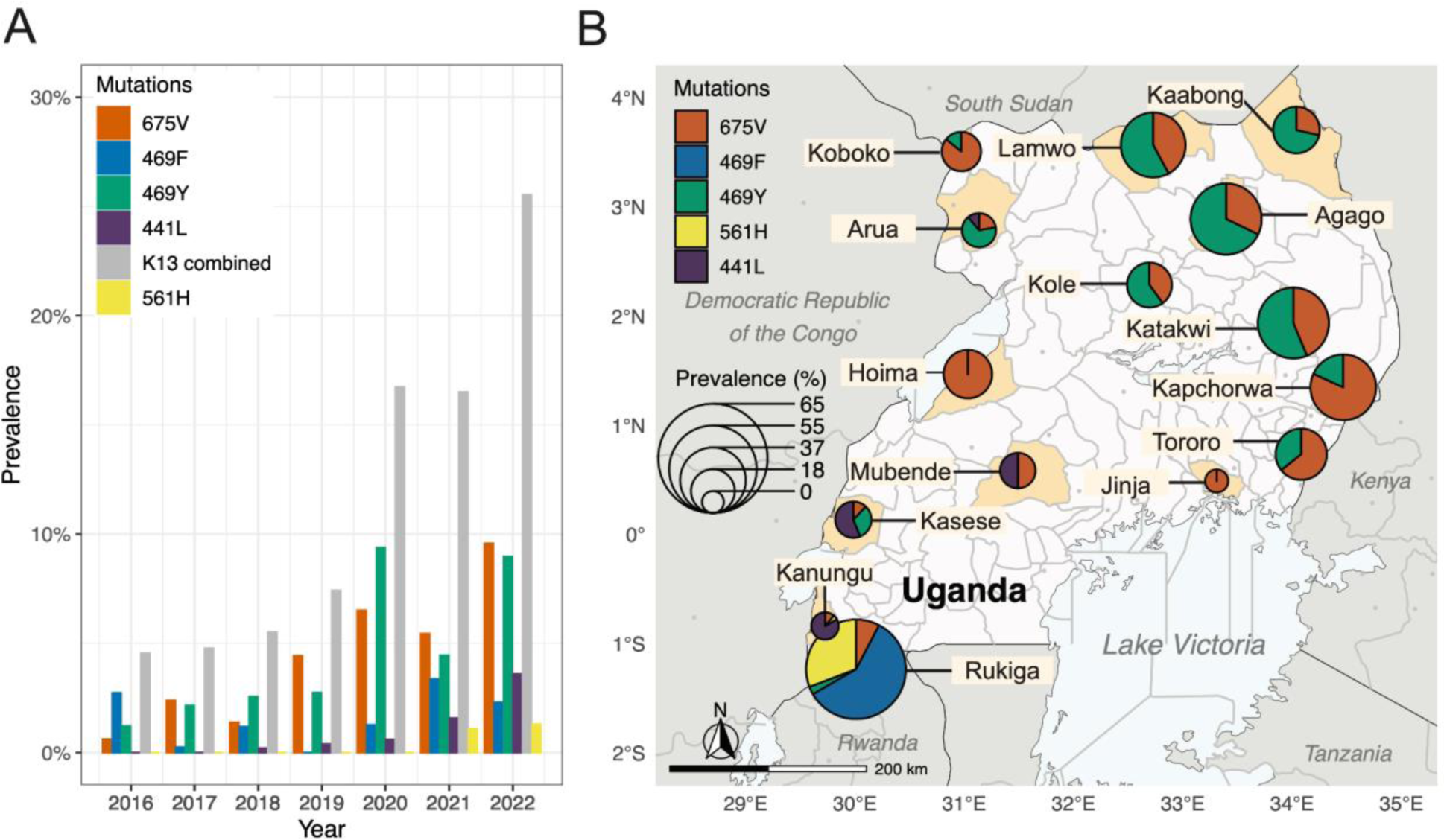
Evolution of ART-R mutation prevalence in Uganda.^20^. (A) Prevalence of the indicated mutations from 2016-2022. (B) The pie-charts represent the distribution and prevalence of mutations in 2022, with study districts shaded and radii of pie charts proportional to the overall prevalence of the five K13 mutations at a site.

Mutation prevalence generally increased over time, but prevalence did not increase uniformly in all districts, with mutation prevalence either not detected consistently or fluctuating over time (Figure 2). Four sites were not included in the subsequent analysis, as they had fewer than three years of positive K13 mutation prevalence (Figure S1). In the northern Ugandan districts, Agago and Lamwo, 469Y increased more quickly than 675V. In the central Ugandan districts, Katakwi and Kole, the increase in prevalence was comparable for 469Y and 675V. 469F was only observed in two western Ugandan districts, Rukiga and Kanungu, with a stronger upward trend observed in Rukiga. In Agago, Kaabong, Kole, and Lamwo, a slight plateau or decrease in 469Y and/or 675V prevalence was observed in 2020-2022. We observed a more consistent trend across all sites when we analyzed the combined K13 mutation prevalence (K13 combined, Figure 2B) than with the individual mutations (Figure 2A).

**Figure 2.**
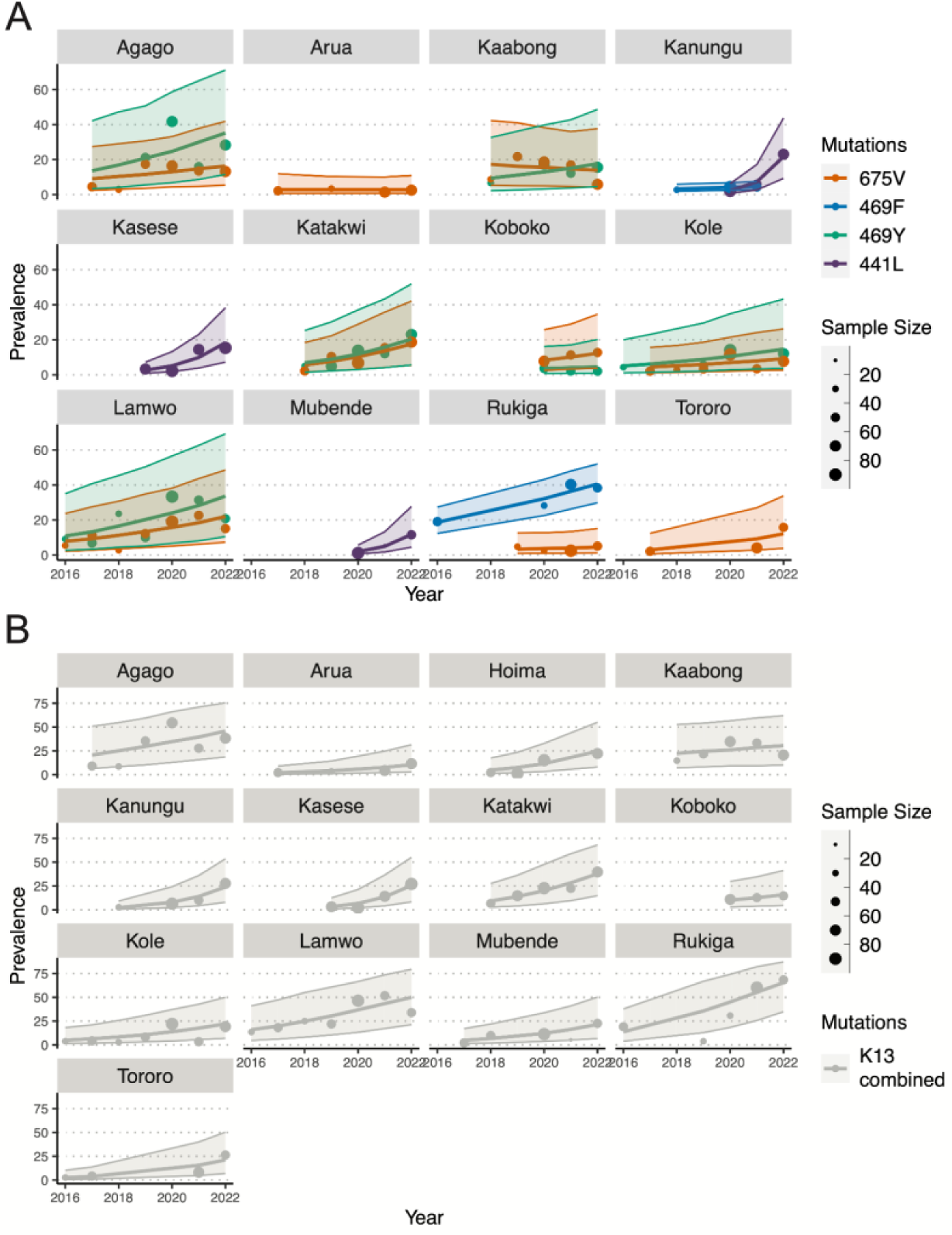
Prevalence trends of K13 mutations across Ugandan sites and model fits. Points indicate the observed prevalence of each genotyped mutation, with sample size indicated by point size. Lines represent the posterior median of Bayesian mixed-effects linear models fitted to the weighted, logit-transformed mutation prevalence data. The shaded areas represent the 95% CrI of the model fit. Panels show the prevalence of the indicated mutations (A) and the prevalence of all five mutations (B, gray, including 561H) in districts with at least three years of non-zero mutation prevalence.

To statistically compare trends in mutation prevalence over time at each site, we modeled the 13 districts with at least three years of non-zero mutation prevalence using Bayesian mixed-effects linear models (Figure 3A, Table S2). We estimate the selection coefficient for all observed K13 mutations combined (including 561H) across all sites at s=0·383 (95% Credible Interval (CrI): 0·247 - 0·528), an average relative increase in prevalence of 38% per year. For individual mutations the selection coefficient was s=0·968 (0·463 - 1·569) for 441L, s=0·153 (−0·445 - 0·727) for 469F, s=0·222 (−0·011 - 0·398) for 469Y, and s=0·152 (−0·023 - 0·312) for 675V (Table S2). While 561H was included in the overall K13 estimates, it was not observed frequently enough to be included in the individual mutation selection coefficient analysis.

**Figure 3.**
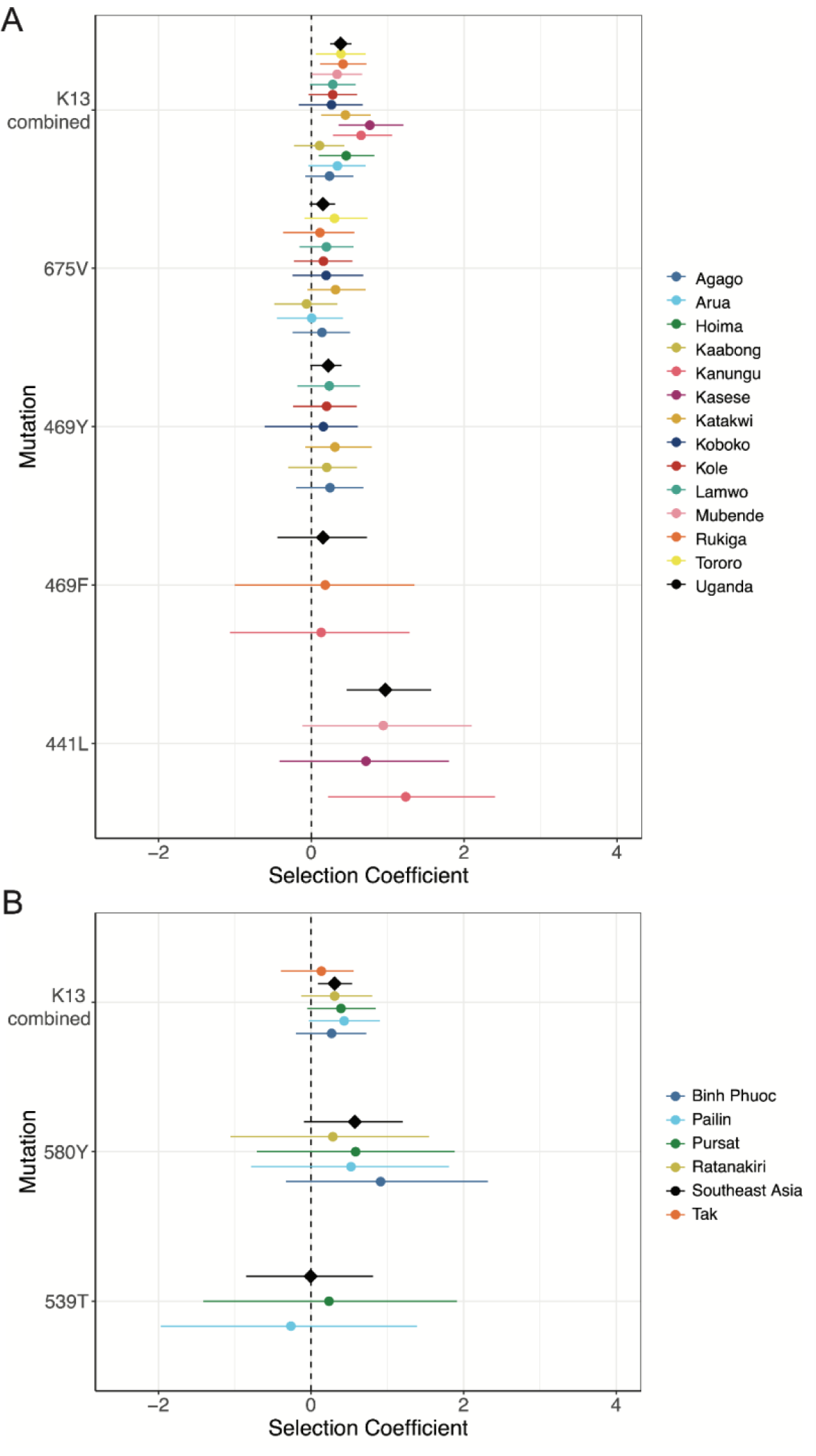
Estimated selection coefficients per year in Uganda and SEA. Per year point estimates for individual sites (colored circles) or all combined sites (black diamonds) are shown with lines representing the 95% CrI for the indicated mutations in Uganda (A) and SEA (B), including the combined mutations (K13 combined).

Selection coefficients were comparable between individual mutations (determined by overlapping 95% CrI) except for 441L, which had a significantly higher selection coefficient. However, data from only three districts informed trends for 441L. The selection was highest for 441L in Kanungu, southwestern Uganda, (s=1·235, 0·215 - 2·404), for 469F (s=0·182, −1·001 - 1·350) in Rukiga, southwestern Uganda, and for 469Y (s=0·309, −0·078 - 0·791) and 675V (s=0·316, −0·053 - 0·721) in Katakwi, northern Uganda. Overall, the selection coefficient for all observed K13 mutations combined (including 561H) was highest in Kasese, southwest Uganda (s=0·765, 0·355 - 1·206) (Table S3).

Lastly, we evaluated how selection in Uganda compared between the first five years (2016-2021) and the full six years (2016-2022) for which data were collected. The selection coefficients across all Ugandan sites for the first five years (2016-2021) were generally slightly higher for individual K13 mutations, with estimates as follows for 441L (s=1·042, 95% CrI: 0·441 - 1·638), 469F (s=0·157, −0·524 - 0·752), 469Y (s=0·361, 0·128 - 0·552), and 675V (s=0·188, −0·029 - 0·358). However, the selection coefficient for all observed K13 mutations combined (including 561H) was lower in the first five years (s=0·360, 0·246 - 0·483) (Table S6, Figure S6).

To compare the spread of ART-R in Uganda and SEA, we estimated selection coefficients during the initial years of spread using the same mixed-effects model (Figure 3B, Figure S4, S5).^23^ Across five districts in three countries in SEA, we estimated a selection coefficient for all observed validated K13 mutations combined of s=0·308 (0·089 - 0·536), and for the two most prevalent validated mutations, 539T and 580Y, of s=−0·005 (−0·852 - 0·814) and s=0·574 (−0·092 - 1·201), respectively. Notably, the selection coefficient for 539T highlights a negative selection, where most of its prevalence time series is positive, but over time, the mutation prevalence declined with a negative selection (Figure S5).

### Ugandan selection coefficients are comparable to previous studies

We identified 79 studies, of which six publications estimated selection coefficients for various antimalarial resistance mutations. These estimates were collated for comparison with selection coefficients estimated in this study (Table S7). The six studies included data from 33 countries collected from 1984-2016. Selection coefficients were previously observed to range between 0·296 and 0.96 (Table S7). The highest and third highest selection coefficients were estimated for the K13 580Y mutation (s=0·960) and various K13 mutations combined (s=0·480), respectively, at the Thailand-Myanmar border from 2001-2014, during the co-emergence of piperaquine resistance in SEA.^21^ The second highest selection coefficient (s=0·660) was estimated at the Thailand-Myanmar border from 1975-1981 for the pyrimethamine resistance mutation PfDHFR 108N.^27^

### Forecasts underpredicts future prevalence in SEA

Comparing the accuracy of forecasts of K13 mutation prevalence based on the initial three to five years of data in SEA, we found that future mutation prevalence was more accurately predicted using all five years of data prevalence dynamics (Figure S7, S8). However, the overall forecasting accuracy was low, with forecasting based on five years of data consistently predicting lower mutation prevalences than those observed in future years, with a weighted mean error of 16% (Table S8). Further, the forecast credible intervals for all validated K13 mutations (Figure 4A) contained only 59%, and for 580Y (Figure 4A) contained 33% of future prevalence points. Despite underestimating future prevalence, the forecast prevalence of K13 580Y still predicted nearing fixation (>95% prevalence) at sites in a median of eight years (range 5-12 years) from initial emergence, which was observed for some sites.

**Figure 4.**
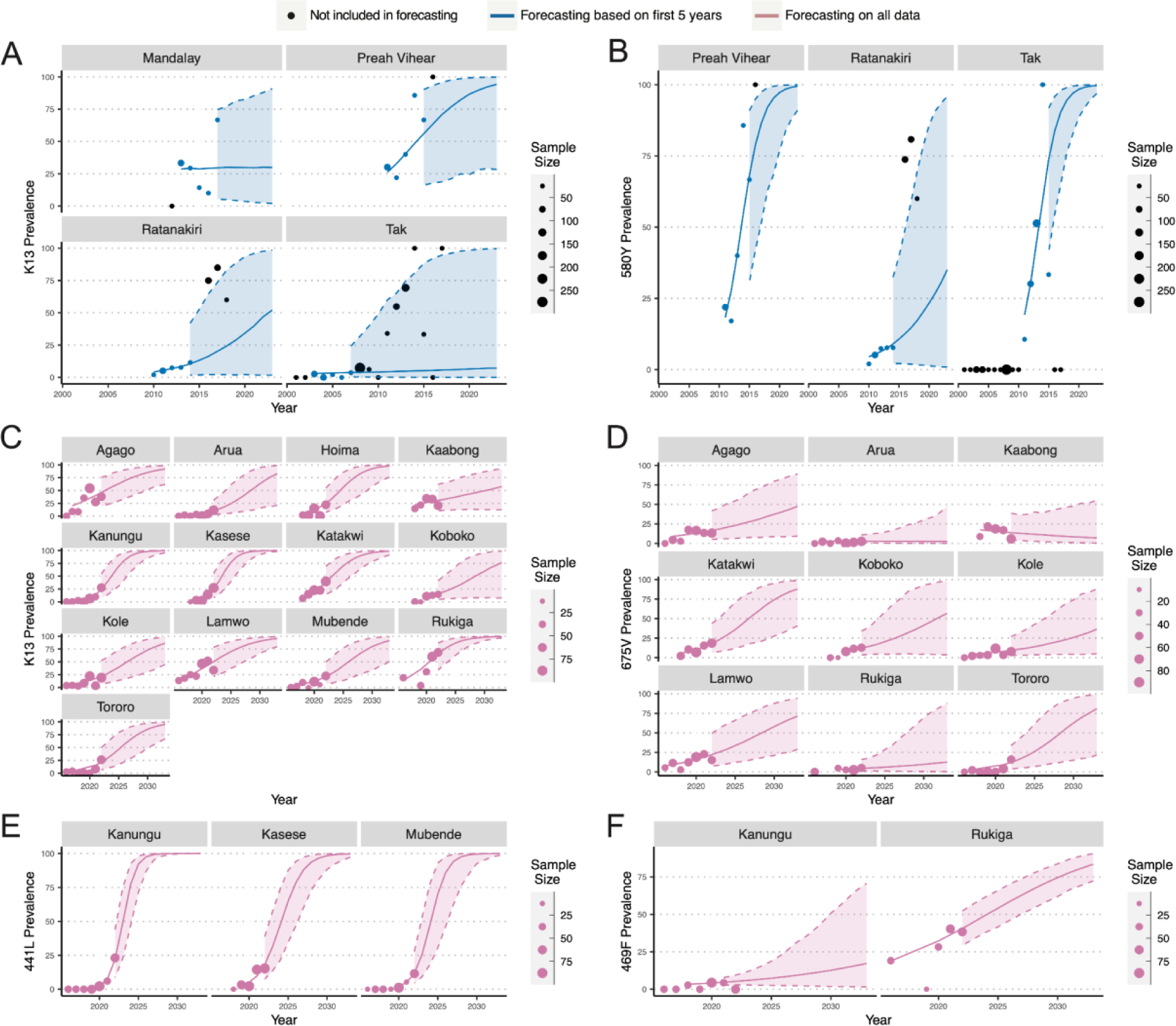
Forecasting ART-R prevalence in SEA and Uganda. Based on the first five years of non-zero prevalence, the prevalence was forecast for all 13 validated K13 markers (A) and 580Y, the most common mutation, in SEA (B). The forecasting was also conducted for all K13 mutations combined (C), 675V (D), 441L (E), and 469F (F) in Uganda. The shaded regions highlight the 95% CrI for the forecast selection.

### Forecast mutation prevalence predicts fixation within a decade in Uganda

We then forecast prevalence in Uganda based on all data for 441L, 469F/Y, and 675V, as well as these K13 mutations combined with 561H (Figure 4C-F). Compared to the forecast prevalence for K13 mutations, the forecasting for individual mutations was more uncertain with wider credible intervals. K13 441L, with the highest selection coefficient, was modeled to near fixation (>95% prevalence) in 2026-2029 in a median of eight years (range six to ten years) after initial emergence. In contrast, 469Y and 675V did not reach fixation with average prevalences of 67% and 44% in 2033, respectively. The forecast combined K13 prevalence across districts was less variable. Seven districts reached near fixation (>95% prevalence) at a median of 12 years (range 9-17 years), leading to a median fixation at the forecast sites in 2030 (2028-2033).

## Discussion

Validated ART-R K13 mutations have increased in prevalence in recent years in eastern Africa, posing a serious threat to malaria control. We estimated the rate of selection in Uganda from 2016-2022 of four key ART-R mutations (441L, 469F/Y, and 675V) and of the combined mutations (441L, 469F/Y, 561H, and 675V). The combined K13 selection coefficient (s=0·383, 95% CrI: 0·247 - 0·528) was comparable to the early spread of K13 mutations in three countries in SEA (s=0·308, 0·089 - 0·536), where artemisinin resistance advanced rapidly a decade ago and quickly became intertwined with ACT partner drug resistance. These findings raise concerns that ACT effectiveness may soon decline in Africa, undermining control efforts and increasing morbidity and mortality.

Three K13 mutations in Uganda, 469F (s=0·153, 95% CrI:−0·445 - 0·727), 469Y (s=0·222, −0·011 - 0·398), and 675V (s=0·152, −0·023 - 0·312), showed lower selection than previously seen in SEA for 580Y (s=0·574, −0·092 - 1·201). In contrast, 441L (s=0·968, 0·463 - 1·569), observed in three districts, which may limit the accuracy of predictions, shows a significantly higher selection coefficient.^20^ In addition, possible plateauing of selection was seen for 469F/Y and 675V, suggesting a decreased magnitude of selection in Uganda. However, the early stages of ART-R selection in SEA were also stochastic, with 580Y only emerging after the initial spread of other K13 mutations, such as 539T.^21^ In future years, additional Ugandan mutations may outcompete those already observed, and our forecasts here for specific mutations may not hold. Despite the potential interplay between mutations, the consistent increase in the prevalence of K13 mutations in Uganda suggests that the overall selection of resistant parasites continues at a concerning pace.

Importantly, the Ugandan malaria setting differs from that in SEA as there is no evidence of prevalent partner drug resistance, which is predicted to potentiate the spread of K13 mutations.^5^ However, this is a growing concern. In particular, decreased susceptibility to lumefantrine, the partner drug in the most widely used ACT, has been observed in ex vivo growth inhibition assays in northern Uganda, and a recent isolate from a UK traveler who returned from Uganda showed multiple recrudescences after treatment with artemether-lumefantrine, with parasites demonstrating a high EC50 to lumefantrine.^15,28^ Also, malaria transmission in Uganda is considerably higher than in previous years after successful transmission reductions through vector control compared to SEA. High transmission is posited to lead to higher host immunity than in low transmission regions, thereby decreasing resistance selection for several reasons. First, with high-level immunity, a greater proportion of infections will be asymptomatic and thus not treated, so a reduced proportion of parasites will be exposed to the drug. Second, infections with relatively unfit drug-resistant parasites are relatively unlikely to cause infections in highly immune individuals. Third, higher transmission intensity increases the proportion of mixed infections, allowing within-host competition between mutant and more-fit wild-type parasites.^5^ Despite these differences, the selection of K13 mutations was comparable between SEA and Uganda, possibly due to effective but interrupted malaria control efforts in Uganda that led to a relatively non-immune population subjected to very high malaria transmission and incidence.^20^

Estimates of future prevalence of drug resistance mutations can predict when antimalarial effectiveness will be lost, informing public health policies and control efforts. In Uganda, the predicted fixation of individual mutations is more variable than combined K13 mutations. For example, K13 441L was predicted to reach near fixation in 2026-2029. However, both 469Y and 675V forecasts were predicted to not fixate within the next ten years, ending with an average prevalence of 67% and 44% in 2033, respectively. Combined K13 mutations are forecast to reach near fixation (>95%) 9-17 years after the mutations were initially observed, leading to a median fixation at the forecast sites in 2030 (2028-2033). These results show that while there is variability between individual mutations across districts, the selection of K13 mutations overall is more consistent across sites. This finding is in agreement with our estimates in SEA, where selection of combined K13 mutation prevalence was more consistent across sites (range equal to 0.136 - 0.434) than that of individual mutations (range equal to −0.263 - 0.912). These findings suggest that the combined selection of all validated K13 mutations may yield more accurate predictions of the risk of ART-R compared to the selection of individual K13 mutations, which have higher variance estimates.

Our analysis has important limitations. First, data on the earliest periods of ART-R emergence in SEA is limited based on retrospective sampling. Second, the accuracy of forecasts made in SEA using only five years of data was notably poor, often underpredicting future mutation prevalence, with the forecast credible intervals for all K13 mutations containing only 59% and for 580Y containing only 33% of future time points. While selection in SEA may have been driven by coincident selection of partner drug (piperaquine) resistance, the selection of K13 mutants in SEA had larger CrI compared to Uganda, which may reflect the differences in sampling schemes between the SEA sites and those in Uganda. Third, our assumption of constant selection for forecast mutation prevalence is oversimplified. Changes in future malaria transmission levels and ACT usage will impact the selection of resistance. Additionally, the importation and geographic spread of resistance is inherently stochastic, particularly in low transmission settings, which makes future resistance dynamics highly uncertain.^29^ Nonetheless, estimates of selection coefficients are useful for parameterizing more sophisticated mechanistic models that explicitly incorporate these factors and can be used to produce scenario forecasts of resistance timelines.^30^

In conclusion, based on our estimates of selection coefficients for ART-R, the rate of increase in K13 mutation prevalence in Uganda (441L, 469F/Y, 561H, 675V) was comparable to the rates estimated for other K13 mutations during the early selection of ART-R in SEA during the late 2000s and early 2010s. Under the assumption of constant selection in Uganda, we predict that the prevalence of all combined K13 mutations will reach near fixation (>95% prevalence) in the majority of sites studied during the next ten years. This information reinforces serious concerns that Africa is facing an extensive increase in ART-R, which may lead to frequent ACT treatment failures if partner drug resistance develops, as occurred in SEA. Continued monitoring of the prevalence of ART-R mutations, both in Uganda and across Africa, combined with studies of the therapeutic efficacy of ACTs, are vitally needed to guide malaria control policy and combat the spread of ART-R and the potential emergence of ACT resistance.

## Contributors

OJW, JAB, and LCO conceived the study. CPGM-S and OJW led the statistical modeling, analysis, and visualization with input from LCO, IG, and DK. VA, TK, SG and MC provided additional data to correctly calculate mutation prevalence from the Uganda data accounting for mixed infections. All authors read, contributed to, and approved the final draft. All authors had full access to all the data in the study and had final responsibility for the decision to submit for publication.

## Data Availability

All data produced are available online at https://github.com/bailey-lab/selmar.

## Acknowledgments

We would specifically like to thank Dominic Kwiatkowski for his help with this manuscript and his supervision of Isaac Ghinai, who started this project. Professor Kwiatkowski sadly passed away while finalizing the manuscript, and we wish to use this space to thank him for his contributions to both this specific study and, more broadly, to the malaria research community. This study, amongst countless others, relies on the openly accessible malaria genome data that MalariaGEN helped to create, and we are immensely grateful to Professor Kwiatkowski for his tireless effort in helping to create this wonderful resource.

## Data Sharing

All our data and code is available on GitHub at bailey-lab/selmar (https://github.com/bailey-lab/selmar).

## Ethics approval and consent to participate

Not applicable.

