## Supplemental Material for "Selection of artemisinin partial resistance Kelch13 mutations in Uganda in 2016-22 was at a rate comparable to that seen previously in South-East Asia"

### Supplementary Materials

### Table of Contents

**Overview of K13 mutation prevalence data in Uganda** **2**
Figure S1: Overview of all prevalence data in Uganda across 16 districts. 2
Table S1: Overview of prevalence data in Uganda 2
Figure S2: Evolution of artemisinin partial resistance in Uganda. 13
**Overview of K13 mutation prevalence data in South-East Asia (SEA)** **14**
Figure S3: Evolution of artemisinin partial resistance mutations in SEA.2 14
Figure S4: Overview of prevalence data in SEA observed at least three times at a given site. 15
**Mixed-effect Bayesian Generalized Linear Model: Estimating selection coefficients in SEA** **16**
Figure S5: Selection of kelch13 mutations in SEA. 16
**Selection Coefficient estimates in Uganda and SEA** **17**
Table S2: Selection coefficients for Uganda 2016-2022. 17
Table S3: Minimum and maximum selection coefficient (s) across Uganda per mutation. 18
Table S4: Selection coefficients for SEA 2003-2018. 18
Table S5: Minimum and maximum selection coefficient (s) across SEA per mutation. 18
**Selection Coefficient estimates in Uganda (2016-2021)** **19**
Table S6: Selection Coefficients for Uganda 2016-2021. 19
Figure S6: Estimated selection coefficients per year in Uganda 2016-2021. 20
**Literature Review** **21**
Table S7: Comparison of selection coefficients obtained from a literature review 21
**Forecasting selection using mixed-effect Bayesian Generalized Linear Model** **22**
Figure S7. Overview of forecasted selection of artemisinin resistance mutations in SEA. 22
Figure S8. Forecasting selection of partial artemisinin resistance mutations in SEA. 23
Table S8: Analysis of forecasting based on the first 3, 4, or 5 years of non-zero prevalence in SEA 23
**References** **24**

### Overview of K13 mutation prevalence data in Uganda

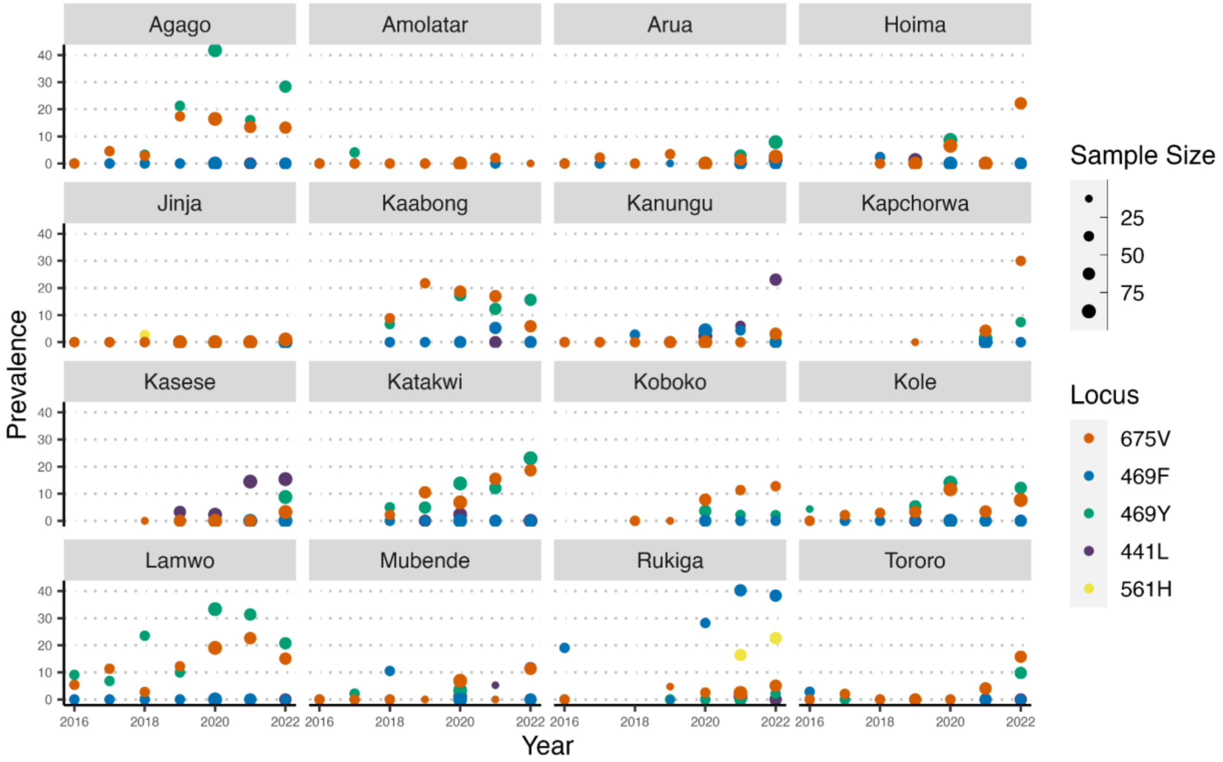


**Figure S1: Overview of all prevalence data in Uganda across 16 districts.**
Prevalence of 675V (red), 469F (blue), 469Y (green), 441L (purple), and 561H (yellow) in Uganda.[1]

**Table S1: Overview of prevalence data in Uganda**

| District | Locus | year | N | X | Prevalence | Min year | N obs | Adj year |
|---|---|---|---|---|---|---|---|---|
| Agago | P441L | 2016 | 30 | 0 | 0 | 2018 | 1 | -2 |
| Agago | P441L | 2017 | 42 | 0 | 0 | 2018 | 1 | -1 |
| Agago | P441L | 2018 | 31 | 1 | 0·03 | 2018 | 1 | 0 |
| Agago | P441L | 2019 | 46 | 0 | 0 | 2018 | 1 | 1 |
| Agago | P441L | 2020 | 79 | 0 | 0 | 2018 | 1 | 2 |
| Agago | P441L | 2021 | 54 | 0 | 0 | 2018 | 1 | 3 |
| Agago | P441L | 2022 | 67 | 0 | 0 | 2018 | 1 | 4 |
| Agago | C469Y | 2016 | 30 | 0 | 0 | 2017 | 6 | -1 |
| Agago | C469Y | 2017 | 44 | 2 | 0·05 | 2017 | 6 | 0 |
| Agago | C469Y | 2018 | 31 | 1 | 0·03 | 2017 | 6 | 1 |
| Agago | C469Y | 2019 | 47 | 10 | 0·21 | 2017 | 6 | 2 |
| Agago | C469Y | 2020 | 79 | 33 | 0·42 | 2017 | 6 | 3 |
| Agago | C469Y | 2021 | 50 | 8 | 0·16 | 2017 | 6 | 4 |
| Agago | C469Y | 2022 | 67 | 19 | 0·28 | 2017 | 6 | 5 |
| Agago | A675V | 2016 | 31 | 0 | 0 | 2017 | 6 | -1 |
| Agago | A675V | 2017 | 44 | 2 | 0·05 | 2017 | 6 | 0 |
| Agago | A675V | 2018 | 35 | 1 | 0·03 | 2017 | 6 | 1 |
| Agago | A675V | 2019 | 46 | 8 | 0·17 | 2017 | 6 | 2 |
| Agago | A675V | 2020 | 79 | 13 | 0·16 | 2017 | 6 | 3 |
| Agago | A675V | 2021 | 52 | 7 | 0·13 | 2017 | 6 | 4 |
| Agago | A675V | 2022 | 68 | 9 | 0·13 | 2017 | 6 | 5 |
| Agago | C469F | 2016 | 30 | 0 | 0 | NA | 0 | NA |
| Agago | C469F | 2017 | 42 | 0 | 0 | NA | 0 | NA |

| | | | | | | | | |
|---|---|---|---|---|---|---|---|---|
| Agago | C469F | 2018 | 30 | 0 | 0 | NA | 0 | NA |
| Agago | C469F | 2019 | 39 | 0 | 0 | NA | 0 | NA |
| Agago | C469F | 2020 | 79 | 0 | 0 | NA | 0 | NA |
| Agago | C469F | 2021 | 50 | 0 | 0 | NA | 0 | NA |
| Agago | C469F | 2022 | 64 | 0 | 0 | NA | 0 | NA |
| Agago | R561H | 2016 | 22 | 0 | 0 | NA | 0 | NA |
| Agago | R561H | 2017 | 44 | 0 | 0 | NA | 0 | NA |
| Agago | R561H | 2018 | 30 | 0 | 0 | NA | 0 | NA |
| Agago | R561H | 2019 | 48 | 0 | 0 | NA | 0 | NA |
| Agago | R561H | 2020 | 79 | 0 | 0 | NA | 0 | NA |
| Agago | R561H | 2021 | 43 | 0 | 0 | NA | 0 | NA |
| Agago | R561H | 2022 | 55 | 0 | 0 | NA | 0 | NA |
| Amolatar | P441L | 2016 | 28 | 0 | 0 | NA | 0 | NA |
| Amolatar | P441L | 2017 | 48 | 0 | 0 | NA | 0 | NA |
| Amolatar | P441L | 2018 | 4 | 0 | 0 | NA | 0 | NA |
| Amolatar | P441L | 2019 | 26 | 0 | 0 | NA | 0 | NA |
| Amolatar | P441L | 2020 | 85 | 0 | 0 | NA | 0 | NA |
| Amolatar | P441L | 2021 | 49 | 0 | 0 | NA | 0 | NA |
| Amolatar | P441L | 2022 | 13 | 0 | 0 | NA | 0 | NA |
| Amolatar | C469Y | 2016 | 28 | 0 | 0 | 2017 | 1 | -1 |
| Amolatar | C469Y | 2017 | 49 | 2 | 0·04 | 2017 | 1 | 0 |
| Amolatar | C469Y | 2018 | 4 | 0 | 0 | 2017 | 1 | 1 |
| Amolatar | C469Y | 2019 | 28 | 0 | 0 | 2017 | 1 | 2 |
| Amolatar | C469Y | 2020 | 85 | 0 | 0 | 2017 | 1 | 3 |
| Amolatar | C469Y | 2021 | 48 | 0 | 0 | 2017 | 1 | 4 |
| Amolatar | C469Y | 2022 | 13 | 0 | 0 | 2017 | 1 | 5 |
| Amolatar | A675V | 2016 | 30 | 0 | 0 | 2021 | 1 | -5 |
| Amolatar | A675V | 2017 | 49 | 0 | 0 | 2021 | 1 | -4 |
| Amolatar | A675V | 2018 | 27 | 0 | 0 | 2021 | 1 | -3 |
| Amolatar | A675V | 2019 | 27 | 0 | 0 | 2021 | 1 | -2 |
| Amolatar | A675V | 2020 | 85 | 0 | 0 | 2021 | 1 | -1 |
| Amolatar | A675V | 2021 | 50 | 1 | 0·02 | 2021 | 1 | 0 |
| Amolatar | A675V | 2022 | 13 | 0 | 0 | 2021 | 1 | 1 |
| Amolatar | C469F | 2016 | 28 | 0 | 0 | NA | 0 | NA |
| Amolatar | C469F | 2017 | 48 | 0 | 0 | NA | 0 | NA |
| Amolatar | C469F | 2018 | 4 | 0 | 0 | NA | 0 | NA |
| Amolatar | C469F | 2019 | 17 | 0 | 0 | NA | 0 | NA |
| Amolatar | C469F | 2020 | 85 | 0 | 0 | NA | 0 | NA |
| Amolatar | C469F | 2021 | 48 | 0 | 0 | NA | 0 | NA |
| Amolatar | C469F | 2022 | 13 | 0 | 0 | NA | 0 | NA |
| Amolatar | R561H | 2016 | 13 | 0 | 0 | NA | 0 | NA |
| Amolatar | R561H | 2017 | 48 | 0 | 0 | NA | 0 | NA |
| Amolatar | R561H | 2018 | 11 | 0 | 0 | NA | 0 | NA |
| Amolatar | R561H | 2019 | 25 | 0 | 0 | NA | 0 | NA |
| Amolatar | R561H | 2020 | 85 | 0 | 0 | NA | 0 | NA |
| Amolatar | R561H | 2021 | 49 | 0 | 0 | NA | 0 | NA |
| Amolatar | R561H | 2022 | 13 | 0 | 0 | NA | 0 | NA |
| Arua | P441L | 2016 | 31 | 0 | 0 | 2022 | 1 | -6 |
| Arua | P441L | 2017 | 42 | 0 | 0 | 2022 | 1 | -5 |
| Arua | P441L | 2018 | 20 | 0 | 0 | 2022 | 1 | -4 |
| Arua | P441L | 2019 | 24 | 0 | 0 | 2022 | 1 | -3 |
| Arua | P441L | 2020 | 81 | 0 | 0 | 2022 | 1 | -2 |
| Arua | P441L | 2021 | 69 | 0 | 0 | 2022 | 1 | -1 |
| Arua | P441L | 2022 | 76 | 1 | 0·01 | 2022 | 1 | 0 |
| Arua | C469Y | 2016 | 31 | 0 | 0 | 2021 | 2 | -5 |

| | | | | | | | | |
|---|---|---|---|---|---|---|---|---|
| Arua | C469Y | 2017 | 45 | 0 | 0 | 2021 | 2 | -4 |
| Arua | C469Y | 2018 | 20 | 0 | 0 | 2021 | 2 | -3 |
| Arua | C469Y | 2019 | 24 | 0 | 0 | 2021 | 2 | -2 |
| Arua | C469Y | 2020 | 81 | 0 | 0 | 2021 | 2 | -1 |
| Arua | C469Y | 2021 | 68 | 2 | 0·03 | 2021 | 2 | 0 |
| Arua | C469Y | 2022 | 76 | 6 | 0·08 | 2021 | 2 | 1 |
| Arua | A675V | 2016 | 33 | 0 | 0 | 2017 | 4 | -1 |
| Arua | A675V | 2017 | 45 | 1 | 0·02 | 2017 | 4 | 0 |
| Arua | A675V | 2018 | 30 | 0 | 0 | 2017 | 4 | 1 |
| Arua | A675V | 2019 | 29 | 1 | 0·03 | 2017 | 4 | 2 |
| Arua | A675V | 2020 | 81 | 0 | 0 | 2017 | 4 | 3 |
| Arua | A675V | 2021 | 68 | 1 | 0·01 | 2017 | 4 | 4 |
| Arua | A675V | 2022 | 78 | 2 | 0·03 | 2017 | 4 | 5 |
| Arua | C469F | 2016 | 31 | 0 | 0 | NA | 0 | NA |
| Arua | C469F | 2017 | 43 | 0 | 0 | NA | 0 | NA |
| Arua | C469F | 2018 | 20 | 0 | 0 | NA | 0 | NA |
| Arua | C469F | 2019 | 10 | 0 | 0 | NA | 0 | NA |
| Arua | C469F | 2020 | 81 | 0 | 0 | NA | 0 | NA |
| Arua | C469F | 2021 | 68 | 0 | 0 | NA | 0 | NA |
| Arua | C469F | 2022 | 70 | 0 | 0 | NA | 0 | NA |
| Arua | R561H | 2016 | 29 | 0 | 0 | NA | 0 | NA |
| Arua | R561H | 2017 | 43 | 0 | 0 | NA | 0 | NA |
| Arua | R561H | 2018 | 24 | 0 | 0 | NA | 0 | NA |
| Arua | R561H | 2019 | 24 | 0 | 0 | NA | 0 | NA |
| Arua | R561H | 2020 | 81 | 0 | 0 | NA | 0 | NA |
| Arua | R561H | 2021 | 56 | 0 | 0 | NA | 0 | NA |
| Arua | R561H | 2022 | 67 | 0 | 0 | NA | 0 | NA |
| Hoima | P441L | 2016 | 0 | 0 | NA | 2019 | 1 | -3 |
| Hoima | P441L | 2017 | 0 | 0 | NA | 2019 | 1 | -2 |
| Hoima | P441L | 2018 | 42 | 0 | 0 | 2019 | 1 | -1 |
| Hoima | P441L | 2019 | 79 | 1 | 0·01 | 2019 | 1 | 0 |
| Hoima | P441L | 2020 | 92 | 0 | 0 | 2019 | 1 | 1 |
| Hoima | P441L | 2021 | 82 | 0 | 0 | 2019 | 1 | 2 |
| Hoima | P441L | 2022 | 60 | 0 | 0 | 2019 | 1 | 3 |
| Hoima | C469Y | 2016 | 0 | 0 | NA | 2020 | 1 | -4 |
| Hoima | C469Y | 2017 | 0 | 0 | NA | 2020 | 1 | -3 |
| Hoima | C469Y | 2018 | 42 | 0 | 0 | 2020 | 1 | -2 |
| Hoima | C469Y | 2019 | 83 | 0 | 0 | 2020 | 1 | -1 |
| Hoima | C469Y | 2020 | 92 | 8 | 0·09 | 2020 | 1 | 0 |
| Hoima | C469Y | 2021 | 82 | 0 | 0 | 2020 | 1 | 1 |
| Hoima | C469Y | 2022 | 60 | 0 | 0 | 2020 | 1 | 2 |
| Hoima | A675V | 2016 | 0 | 0 | NA | 2020 | 2 | -4 |
| Hoima | A675V | 2017 | 0 | 0 | NA | 2020 | 2 | -3 |
| Hoima | A675V | 2018 | 45 | 0 | 0 | 2020 | 2 | -2 |
| Hoima | A675V | 2019 | 82 | 0 | 0 | 2020 | 2 | -1 |
| Hoima | A675V | 2020 | 92 | 6 | 0·07 | 2020 | 2 | 0 |
| Hoima | A675V | 2021 | 82 | 0 | 0 | 2020 | 2 | 1 |
| Hoima | A675V | 2022 | 63 | 14 | 0·22 | 2020 | 2 | 2 |
| Hoima | C469F | 2016 | 0 | 0 | NA | 2018 | 1 | -2 |
| Hoima | C469F | 2017 | 0 | 0 | NA | 2018 | 1 | -1 |
| Hoima | C469F | 2018 | 42 | 1 | 0·02 | 2018 | 1 | 0 |
| Hoima | C469F | 2019 | 47 | 0 | 0 | 2018 | 1 | 1 |
| Hoima | C469F | 2020 | 92 | 0 | 0 | 2018 | 1 | 2 |
| Hoima | C469F | 2021 | 82 | 0 | 0 | 2018 | 1 | 3 |
| Hoima | C469F | 2022 | 60 | 0 | 0 | 2018 | 1 | 4 |

| | | | | | | | | |
|---|---|---|---|---|---|---|---|---|
| Hoima | R561H | 2016 | 0 | 0 | NA | NA | 0 | NA |
| Hoima | R561H | 2017 | 0 | 0 | NA | NA | 0 | NA |
| Hoima | R561H | 2018 | 42 | 0 | 0 | NA | 0 | NA |
| Hoima | R561H | 2019 | 82 | 0 | 0 | NA | 0 | NA |
| Hoima | R561H | 2020 | 92 | 0 | 0 | NA | 0 | NA |
| Hoima | R561H | 2021 | 65 | 0 | 0 | NA | 0 | NA |
| Hoima | R561H | 2022 | 53 | 0 | 0 | NA | 0 | NA |
| Jinja | P441L | 2016 | 29 | 0 | 0 | NA | 0 | NA |
| Jinja | P441L | 2017 | 48 | 0 | 0 | NA | 0 | NA |
| Jinja | P441L | 2018 | 37 | 0 | 0 | NA | 0 | NA |
| Jinja | P441L | 2019 | 89 | 0 | 0 | NA | 0 | NA |
| Jinja | P441L | 2020 | 96 | 0 | 0 | NA | 0 | NA |
| Jinja | P441L | 2021 | 95 | 0 | 0 | NA | 0 | NA |
| Jinja | P441L | 2022 | 99 | 0 | 0 | NA | 0 | NA |
| Jinja | C469Y | 2016 | 29 | 0 | 0 | NA | 0 | NA |
| Jinja | C469Y | 2017 | 48 | 0 | 0 | NA | 0 | NA |
| Jinja | C469Y | 2018 | 37 | 0 | 0 | NA | 0 | NA |
| Jinja | C469Y | 2019 | 91 | 0 | 0 | NA | 0 | NA |
| Jinja | C469Y | 2020 | 96 | 0 | 0 | NA | 0 | NA |
| Jinja | C469Y | 2021 | 95 | 0 | 0 | NA | 0 | NA |
| Jinja | C469Y | 2022 | 99 | 0 | 0 | NA | 0 | NA |
| Jinja | A675V | 2016 | 29 | 0 | 0 | 2022 | 1 | -6 |
| Jinja | A675V | 2017 | 48 | 0 | 0 | 2022 | 1 | -5 |
| Jinja | A675V | 2018 | 40 | 0 | 0 | 2022 | 1 | -4 |
| Jinja | A675V | 2019 | 90 | 0 | 0 | 2022 | 1 | -3 |
| Jinja | A675V | 2020 | 96 | 0 | 0 | 2022 | 1 | -2 |
| Jinja | A675V | 2021 | 95 | 0 | 0 | 2022 | 1 | -1 |
| Jinja | A675V | 2022 | 99 | 1 | 0·01 | 2022 | 1 | 0 |
| Jinja | C469F | 2016 | 29 | 0 | 0 | NA | 0 | NA |
| Jinja | C469F | 2017 | 48 | 0 | 0 | NA | 0 | NA |
| Jinja | C469F | 2018 | 37 | 0 | 0 | NA | 0 | NA |
| Jinja | C469F | 2019 | 43 | 0 | 0 | NA | 0 | NA |
| Jinja | C469F | 2020 | 96 | 0 | 0 | NA | 0 | NA |
| Jinja | C469F | 2021 | 95 | 0 | 0 | NA | 0 | NA |
| Jinja | C469F | 2022 | 99 | 0 | 0 | NA | 0 | NA |
| Jinja | R561H | 2016 | 27 | 0 | 0 | 2018 | 1 | -2 |
| Jinja | R561H | 2017 | 48 | 0 | 0 | 2018 | 1 | -1 |
| Jinja | R561H | 2018 | 38 | 1 | 0·03 | 2018 | 1 | 0 |
| Jinja | R561H | 2019 | 89 | 0 | 0 | 2018 | 1 | 1 |
| Jinja | R561H | 2020 | 96 | 0 | 0 | 2018 | 1 | 2 |
| Jinja | R561H | 2021 | 74 | 0 | 0 | 2018 | 1 | 3 |
| Jinja | R561H | 2022 | 92 | 0 | 0 | 2018 | 1 | 4 |
| Kaabong | P441L | 2016 | 0 | 0 | NA | NA | 0 | NA |
| Kaabong | P441L | 2017 | 0 | 0 | NA | NA | 0 | NA |
| Kaabong | P441L | 2018 | 30 | 0 | 0 | NA | 0 | NA |
| Kaabong | P441L | 2019 | 45 | 0 | 0 | NA | 0 | NA |
| Kaabong | P441L | 2020 | 75 | 0 | 0 | NA | 0 | NA |
| Kaabong | P441L | 2021 | 60 | 0 | 0 | NA | 0 | NA |
| Kaabong | P441L | 2022 | 64 | 0 | 0 | NA | 0 | NA |
| Kaabong | C469Y | 2016 | 0 | 0 | NA | 2018 | 4 | -2 |
| Kaabong | C469Y | 2017 | 0 | 0 | NA | 2018 | 4 | -1 |
| Kaabong | C469Y | 2018 | 30 | 2 | 0·07 | 2018 | 4 | 0 |
| Kaabong | C469Y | 2019 | 45 | 0 | 0 | 2018 | 4 | 1 |
| Kaabong | C469Y | 2020 | 75 | 13 | 0·17 | 2018 | 4 | 2 |
| Kaabong | C469Y | 2021 | 57 | 7 | 0·12 | 2018 | 4 | 3 |

| | | | | | | | | |
|---|---|---|---|---|---|---|---|---|
| Kaabong | C469Y | 2022 | 64 | 10 | 0·16 | 2018 | 4 | 4 |
| Kaabong | A675V | 2016 | 0 | 0 | NA | 2018 | 5 | -2 |
| Kaabong | A675V | 2017 | 0 | 0 | NA | 2018 | 5 | -1 |
| Kaabong | A675V | 2018 | 34 | 3 | 0·09 | 2018 | 5 | 0 |
| Kaabong | A675V | 2019 | 46 | 10 | 0·22 | 2018 | 5 | 1 |
| Kaabong | A675V | 2020 | 75 | 14 | 0·19 | 2018 | 5 | 2 |
| Kaabong | A675V | 2021 | 59 | 10 | 0·17 | 2018 | 5 | 3 |
| Kaabong | A675V | 2022 | 68 | 4 | 0·06 | 2018 | 5 | 4 |
| Kaabong | C469F | 2016 | 0 | 0 | NA | 2021 | 1 | -5 |
| Kaabong | C469F | 2017 | 0 | 0 | NA | 2021 | 1 | -4 |
| Kaabong | C469F | 2018 | 29 | 0 | 0 | 2021 | 1 | -3 |
| Kaabong | C469F | 2019 | 31 | 0 | 0 | 2021 | 1 | -2 |
| Kaabong | C469F | 2020 | 75 | 0 | 0 | 2021 | 1 | -1 |
| Kaabong | C469F | 2021 | 57 | 3 | 0·05 | 2021 | 1 | 0 |
| Kaabong | C469F | 2022 | 60 | 0 | 0 | 2021 | 1 | 1 |
| Kaabong | R561H | 2016 | 0 | 0 | NA | NA | 0 | NA |
| Kaabong | R561H | 2017 | 0 | 0 | NA | NA | 0 | NA |
| Kaabong | R561H | 2018 | 30 | 0 | 0 | NA | 0 | NA |
| Kaabong | R561H | 2019 | 44 | 0 | 0 | NA | 0 | NA |
| Kaabong | R561H | 2020 | 75 | 0 | 0 | NA | 0 | NA |
| Kaabong | R561H | 2021 | 40 | 0 | 0 | NA | 0 | NA |
| Kaabong | R561H | 2022 | 54 | 0 | 0 | NA | 0 | NA |
| Kanungu | P441L | 2016 | 37 | 0 | 0 | 2020 | 3 | -4 |
| Kanungu | P441L | 2017 | 48 | 0 | 0 | 2020 | 3 | -3 |
| Kanungu | P441L | 2018 | 36 | 0 | 0 | 2020 | 3 | -2 |
| Kanungu | P441L | 2019 | 59 | 0 | 0 | 2020 | 3 | -1 |
| Kanungu | P441L | 2020 | 91 | 2 | 0·02 | 2020 | 3 | 0 |
| Kanungu | P441L | 2021 | 50 | 3 | 0·06 | 2020 | 3 | 1 |
| Kanungu | P441L | 2022 | 65 | 15 | 0·23 | 2020 | 3 | 2 |
| Kanungu | C469Y | 2016 | 37 | 0 | 0 | NA | 0 | NA |
| Kanungu | C469Y | 2017 | 48 | 0 | 0 | NA | 0 | NA |
| Kanungu | C469Y | 2018 | 35 | 0 | 0 | NA | 0 | NA |
| Kanungu | C469Y | 2019 | 64 | 0 | 0 | NA | 0 | NA |
| Kanungu | C469Y | 2020 | 91 | 0 | 0 | NA | 0 | NA |
| Kanungu | C469Y | 2021 | 46 | 0 | 0 | NA | 0 | NA |
| Kanungu | C469Y | 2022 | 65 | 0 | 0 | NA | 0 | NA |
| Kanungu | A675V | 2016 | 37 | 0 | 0 | 2022 | 1 | -6 |
| Kanungu | A675V | 2017 | 48 | 0 | 0 | 2022 | 1 | -5 |
| Kanungu | A675V | 2018 | 38 | 0 | 0 | 2022 | 1 | -4 |
| Kanungu | A675V | 2019 | 61 | 0 | 0 | 2022 | 1 | -3 |
| Kanungu | A675V | 2020 | 94 | 0 | 0 | 2022 | 1 | -2 |
| Kanungu | A675V | 2021 | 44 | 0 | 0 | 2022 | 1 | -1 |
| Kanungu | A675V | 2022 | 64 | 2 | 0·03 | 2022 | 1 | 0 |
| Kanungu | C469F | 2016 | 37 | 0 | 0 | 2018 | 3 | -2 |
| Kanungu | C469F | 2017 | 48 | 0 | 0 | 2018 | 3 | -1 |
| Kanungu | C469F | 2018 | 36 | 1 | 0·03 | 2018 | 3 | 0 |
| Kanungu | C469F | 2019 | 40 | 0 | 0 | 2018 | 3 | 1 |
| Kanungu | C469F | 2020 | 91 | 4 | 0·04 | 2018 | 3 | 2 |
| Kanungu | C469F | 2021 | 46 | 2 | 0·04 | 2018 | 3 | 3 |
| Kanungu | C469F | 2022 | 65 | 0 | 0 | 2018 | 3 | 4 |
| Kanungu | R561H | 2016 | 37 | 0 | 0 | 2022 | 1 | -6 |
| Kanungu | R561H | 2017 | 48 | 0 | 0 | 2022 | 1 | -5 |
| Kanungu | R561H | 2018 | 36 | 0 | 0 | 2022 | 1 | -4 |
| Kanungu | R561H | 2019 | 62 | 0 | 0 | 2022 | 1 | -3 |
| Kanungu | R561H | 2020 | 91 | 0 | 0 | 2022 | 1 | -2 |

| | | | | | | | | |
|---|---|---|---|---|---|---|---|---|
| Kanungu | R561H | 2021 | 37 | 0 | 0 | 2022 | 1 | -1 |
| Kanungu | R561H | 2022 | 51 | 1 | 0·02 | 2022 | 1 | 0 |
| Kapchorwa | P441L | 2016 | 0 | 0 | NA | NA | 0 | NA |
| Kapchorwa | P441L | 2017 | 0 | 0 | NA | NA | 0 | NA |
| Kapchorwa | P441L | 2018 | 0 | 0 | NA | NA | 0 | NA |
| Kapchorwa | P441L | 2019 | 16 | 0 | 0 | NA | 0 | NA |
| Kapchorwa | P441L | 2020 | 0 | 0 | NA | NA | 0 | NA |
| Kapchorwa | P441L | 2021 | 77 | 0 | 0 | NA | 0 | NA |
| Kapchorwa | P441L | 2022 | 27 | 0 | 0 | NA | 0 | NA |
| Kapchorwa | C469Y | 2016 | 0 | 0 | NA | 2021 | 2 | -5 |
| Kapchorwa | C469Y | 2017 | 0 | 0 | NA | 2021 | 2 | -4 |
| Kapchorwa | C469Y | 2018 | 0 | 0 | NA | 2021 | 2 | -3 |
| Kapchorwa | C469Y | 2019 | 16 | 0 | 0 | 2021 | 2 | -2 |
| Kapchorwa | C469Y | 2020 | 0 | 0 | NA | 2021 | 2 | -1 |
| Kapchorwa | C469Y | 2021 | 77 | 1 | 0·01 | 2021 | 2 | 0 |
| Kapchorwa | C469Y | 2022 | 27 | 2 | 0·07 | 2021 | 2 | 1 |
| Kapchorwa | A675V | 2016 | 0 | 0 | NA | 2021 | 2 | -5 |
| Kapchorwa | A675V | 2017 | 0 | 0 | NA | 2021 | 2 | -4 |
| Kapchorwa | A675V | 2018 | 0 | 0 | NA | 2021 | 2 | -3 |
| Kapchorwa | A675V | 2019 | 15 | 0 | 0 | 2021 | 2 | -2 |
| Kapchorwa | A675V | 2020 | 0 | 0 | NA | 2021 | 2 | -1 |
| Kapchorwa | A675V | 2021 | 71 | 3 | 0·04 | 2021 | 2 | 0 |
| Kapchorwa | A675V | 2022 | 30 | 9 | 0·3 | 2021 | 2 | 1 |
| Kapchorwa | C469F | 2016 | 0 | 0 | NA | NA | 0 | NA |
| Kapchorwa | C469F | 2017 | 0 | 0 | NA | NA | 0 | NA |
| Kapchorwa | C469F | 2018 | 0 | 0 | NA | NA | 0 | NA |
| Kapchorwa | C469F | 2019 | 16 | 0 | 0 | NA | 0 | NA |
| Kapchorwa | C469F | 2020 | 0 | 0 | NA | NA | 0 | NA |
| Kapchorwa | C469F | 2021 | 77 | 0 | 0 | NA | 0 | NA |
| Kapchorwa | C469F | 2022 | 26 | 0 | 0 | NA | 0 | NA |
| Kapchorwa | R561H | 2016 | 0 | 0 | NA | NA | 0 | NA |
| Kapchorwa | R561H | 2017 | 0 | 0 | NA | NA | 0 | NA |
| Kapchorwa | R561H | 2018 | 0 | 0 | NA | NA | 0 | NA |
| Kapchorwa | R561H | 2019 | 14 | 0 | 0 | NA | 0 | NA |
| Kapchorwa | R561H | 2020 | 0 | 0 | NA | NA | 0 | NA |
| Kapchorwa | R561H | 2021 | 69 | 0 | 0 | NA | 0 | NA |
| Kapchorwa | R561H | 2022 | 12 | 0 | 0 | NA | 0 | NA |
| Kasese | P441L | 2016 | 0 | 0 | NA | 2019 | 4 | -3 |
| Kasese | P441L | 2017 | 0 | 0 | NA | 2019 | 4 | -2 |
| Kasese | P441L | 2018 | 19 | 0 | 0 | 2019 | 4 | -1 |
| Kasese | P441L | 2019 | 61 | 2 | 0·03 | 2019 | 4 | 0 |
| Kasese | P441L | 2020 | 89 | 2 | 0·02 | 2019 | 4 | 1 |
| Kasese | P441L | 2021 | 76 | 11 | 0·14 | 2019 | 4 | 2 |
| Kasese | P441L | 2022 | 91 | 14 | 0·15 | 2019 | 4 | 3 |
| Kasese | C469Y | 2016 | 0 | 0 | NA | 2022 | 1 | -6 |
| Kasese | C469Y | 2017 | 0 | 0 | NA | 2022 | 1 | -5 |
| Kasese | C469Y | 2018 | 19 | 0 | 0 | 2022 | 1 | -4 |
| Kasese | C469Y | 2019 | 61 | 0 | 0 | 2022 | 1 | -3 |
| Kasese | C469Y | 2020 | 89 | 0 | 0 | 2022 | 1 | -2 |
| Kasese | C469Y | 2021 | 78 | 0 | 0 | 2022 | 1 | -1 |
| Kasese | C469Y | 2022 | 91 | 8 | 0·09 | 2022 | 1 | 0 |
| Kasese | A675V | 2016 | 0 | 0 | NA | 2022 | 1 | -6 |
| Kasese | A675V | 2017 | 0 | 0 | NA | 2022 | 1 | -5 |
| Kasese | A675V | 2018 | 24 | 0 | 0 | 2022 | 1 | -4 |
| Kasese | A675V | 2019 | 64 | 0 | 0 | 2022 | 1 | -3 |

| | | | | | | | | |
|---|---|---|---|---|---|---|---|---|
| Kasese | A675V | 2020 | 92 | 0 | 0 | 2022 | 1 | -2 |
| Kasese | A675V | 2021 | 75 | 0 | 0 | 2022 | 1 | -1 |
| Kasese | A675V | 2022 | 92 | 3 | 0·03 | 2022 | 1 | 0 |
| Kasese | C469F | 2016 | 0 | 0 | NA | NA | 0 | NA |
| Kasese | C469F | 2017 | 0 | 0 | NA | NA | 0 | NA |
| Kasese | C469F | 2018 | 19 | 0 | 0 | NA | 0 | NA |
| Kasese | C469F | 2019 | 39 | 0 | 0 | NA | 0 | NA |
| Kasese | C469F | 2020 | 89 | 0 | 0 | NA | 0 | NA |
| Kasese | C469F | 2021 | 78 | 0 | 0 | NA | 0 | NA |
| Kasese | C469F | 2022 | 88 | 0 | 0 | NA | 0 | NA |
| Kasese | R561H | 2016 | 0 | 0 | NA | NA | 0 | NA |
| Kasese | R561H | 2017 | 0 | 0 | NA | NA | 0 | NA |
| Kasese | R561H | 2018 | 17 | 0 | 0 | NA | 0 | NA |
| Kasese | R561H | 2019 | 60 | 0 | 0 | NA | 0 | NA |
| Kasese | R561H | 2020 | 90 | 0 | 0 | NA | 0 | NA |
| Kasese | R561H | 2021 | 61 | 0 | 0 | NA | 0 | NA |
| Kasese | R561H | 2022 | 85 | 0 | 0 | NA | 0 | NA |
| Katakwi | P441L | 2016 | 0 | 0 | NA | 2020 | 1 | -4 |
| Katakwi | P441L | 2017 | 0 | 0 | NA | 2020 | 1 | -3 |
| Katakwi | P441L | 2018 | 40 | 0 | 0 | 2020 | 1 | -2 |
| Katakwi | P441L | 2019 | 55 | 0 | 0 | 2020 | 1 | -1 |
| Katakwi | P441L | 2020 | 87 | 2 | 0·02 | 2020 | 1 | 0 |
| Katakwi | P441L | 2021 | 58 | 0 | 0 | 2020 | 1 | 1 |
| Katakwi | P441L | 2022 | 78 | 0 | 0 | 2020 | 1 | 2 |
| Katakwi | C469Y | 2016 | 0 | 0 | NA | 2018 | 5 | -2 |
| Katakwi | C469Y | 2017 | 0 | 0 | NA | 2018 | 5 | -1 |
| Katakwi | C469Y | 2018 | 40 | 2 | 0·05 | 2018 | 5 | 0 |
| Katakwi | C469Y | 2019 | 61 | 3 | 0·05 | 2018 | 5 | 1 |
| Katakwi | C469Y | 2020 | 87 | 12 | 0·14 | 2018 | 5 | 2 |
| Katakwi | C469Y | 2021 | 58 | 7 | 0·12 | 2018 | 5 | 3 |
| Katakwi | C469Y | 2022 | 78 | 18 | 0·23 | 2018 | 5 | 4 |
| Katakwi | A675V | 2016 | 0 | 0 | NA | 2018 | 5 | -2 |
| Katakwi | A675V | 2017 | 0 | 0 | NA | 2018 | 5 | -1 |
| Katakwi | A675V | 2018 | 45 | 1 | 0·02 | 2018 | 5 | 0 |
| Katakwi | A675V | 2019 | 57 | 6 | 0·11 | 2018 | 5 | 1 |
| Katakwi | A675V | 2020 | 88 | 6 | 0·07 | 2018 | 5 | 2 |
| Katakwi | A675V | 2021 | 58 | 9 | 0·16 | 2018 | 5 | 3 |
| Katakwi | A675V | 2022 | 75 | 14 | 0·19 | 2018 | 5 | 4 |
| Katakwi | C469F | 2016 | 0 | 0 | NA | NA | 0 | NA |
| Katakwi | C469F | 2017 | 0 | 0 | NA | NA | 0 | NA |
| Katakwi | C469F | 2018 | 40 | 0 | 0 | NA | 0 | NA |
| Katakwi | C469F | 2019 | 36 | 0 | 0 | NA | 0 | NA |
| Katakwi | C469F | 2020 | 86 | 0 | 0 | NA | 0 | NA |
| Katakwi | C469F | 2021 | 58 | 0 | 0 | NA | 0 | NA |
| Katakwi | C469F | 2022 | 68 | 0 | 0 | NA | 0 | NA |
| Katakwi | R561H | 2016 | 0 | 0 | NA | NA | 0 | NA |
| Katakwi | R561H | 2017 | 0 | 0 | NA | NA | 0 | NA |
| Katakwi | R561H | 2018 | 40 | 0 | 0 | NA | 0 | NA |
| Katakwi | R561H | 2019 | 57 | 0 | 0 | NA | 0 | NA |
| Katakwi | R561H | 2020 | 86 | 0 | 0 | NA | 0 | NA |
| Katakwi | R561H | 2021 | 41 | 0 | 0 | NA | 0 | NA |
| Katakwi | R561H | 2022 | 62 | 0 | 0 | NA | 0 | NA |
| Koboko | P441L | 2016 | 0 | 0 | NA | NA | 0 | NA |
| Koboko | P441L | 2017 | 0 | 0 | NA | NA | 0 | NA |
| Koboko | P441L | 2018 | 23 | 0 | 0 | NA | 0 | NA |

| | | | | | | | | |
|---|---|---|---|---|---|---|---|---|
| Koboko | P441L | 2019 | 4 | 0 | 0 | NA | 0 | NA |
| Koboko | P441L | 2020 | 55 | 0 | 0 | NA | 0 | NA |
| Koboko | P441L | 2021 | 47 | 0 | 0 | NA | 0 | NA |
| Koboko | P441L | 2022 | 48 | 0 | 0 | NA | 0 | NA |
| Koboko | C469Y | 2016 | 0 | 0 | NA | 2020 | 3 | -4 |
| Koboko | C469Y | 2017 | 0 | 0 | NA | 2020 | 3 | -3 |
| Koboko | C469Y | 2018 | 23 | 0 | 0 | 2020 | 3 | -2 |
| Koboko | C469Y | 2019 | 4 | 0 | 0 | 2020 | 3 | -1 |
| Koboko | C469Y | 2020 | 55 | 2 | 0·04 | 2020 | 3 | 0 |
| Koboko | C469Y | 2021 | 46 | 1 | 0·02 | 2020 | 3 | 1 |
| Koboko | C469Y | 2022 | 48 | 1 | 0·02 | 2020 | 3 | 2 |
| Koboko | A675V | 2016 | 0 | 0 | NA | 2020 | 3 | -4 |
| Koboko | A675V | 2017 | 0 | 0 | NA | 2020 | 3 | -3 |
| Koboko | A675V | 2018 | 34 | 0 | 0 | 2020 | 3 | -2 |
| Koboko | A675V | 2019 | 5 | 0 | 0 | 2020 | 3 | -1 |
| Koboko | A675V | 2020 | 64 | 5 | 0·08 | 2020 | 3 | 0 |
| Koboko | A675V | 2021 | 44 | 5 | 0·11 | 2020 | 3 | 1 |
| Koboko | A675V | 2022 | 47 | 6 | 0·13 | 2020 | 3 | 2 |
| Koboko | C469F | 2016 | 0 | 0 | NA | NA | 0 | NA |
| Koboko | C469F | 2017 | 0 | 0 | NA | NA | 0 | NA |
| Koboko | C469F | 2018 | 23 | 0 | 0 | NA | 0 | NA |
| Koboko | C469F | 2019 | 4 | 0 | 0 | NA | 0 | NA |
| Koboko | C469F | 2020 | 54 | 0 | 0 | NA | 0 | NA |
| Koboko | C469F | 2021 | 46 | 0 | 0 | NA | 0 | NA |
| Koboko | C469F | 2022 | 47 | 0 | 0 | NA | 0 | NA |
| Koboko | R561H | 2016 | 0 | 0 | NA | NA | 0 | NA |
| Koboko | R561H | 2017 | 0 | 0 | NA | NA | 0 | NA |
| Koboko | R561H | 2018 | 26 | 0 | 0 | NA | 0 | NA |
| Koboko | R561H | 2019 | 3 | 0 | 0 | NA | 0 | NA |
| Koboko | R561H | 2020 | 62 | 0 | 0 | NA | 0 | NA |
| Koboko | R561H | 2021 | 27 | 0 | 0 | NA | 0 | NA |
| Koboko | R561H | 2022 | 35 | 0 | 0 | NA | 0 | NA |
| Kole | P441L | 2016 | 23 | 0 | 0 | NA | 0 | NA |
| Kole | P441L | 2017 | 46 | 0 | 0 | NA | 0 | NA |
| Kole | P441L | 2018 | 36 | 0 | 0 | NA | 0 | NA |
| Kole | P441L | 2019 | 56 | 0 | 0 | NA | 0 | NA |
| Kole | P441L | 2020 | 85 | 0 | 0 | NA | 0 | NA |
| Kole | P441L | 2021 | 56 | 0 | 0 | NA | 0 | NA |
| Kole | P441L | 2022 | 74 | 0 | 0 | NA | 0 | NA |
| Kole | C469Y | 2016 | 23 | 1 | 0·04 | 2016 | 5 | 0 |
| Kole | C469Y | 2017 | 47 | 1 | 0·02 | 2016 | 5 | 1 |
| Kole | C469Y | 2018 | 36 | 0 | 0 | 2016 | 5 | 2 |
| Kole | C469Y | 2019 | 56 | 3 | 0·05 | 2016 | 5 | 3 |
| Kole | C469Y | 2020 | 85 | 12 | 0·14 | 2016 | 5 | 4 |
| Kole | C469Y | 2021 | 54 | 0 | 0 | 2016 | 5 | 5 |
| Kole | C469Y | 2022 | 74 | 9 | 0·12 | 2016 | 5 | 6 |
| Kole | A675V | 2016 | 26 | 0 | 0 | 2017 | 6 | -1 |
| Kole | A675V | 2017 | 47 | 1 | 0·02 | 2017 | 6 | 0 |
| Kole | A675V | 2018 | 34 | 1 | 0·03 | 2017 | 6 | 1 |
| Kole | A675V | 2019 | 60 | 2 | 0·03 | 2017 | 6 | 2 |
| Kole | A675V | 2020 | 86 | 10 | 0·12 | 2017 | 6 | 3 |
| Kole | A675V | 2021 | 58 | 2 | 0·03 | 2017 | 6 | 4 |
| Kole | A675V | 2022 | 78 | 6 | 0·08 | 2017 | 6 | 5 |
| Kole | C469F | 2016 | 23 | 0 | 0 | NA | 0 | NA |
| Kole | C469F | 2017 | 47 | 0 | 0 | NA | 0 | NA |

| | | | | | | | | |
|---|---|---|---|---|---|---|---|---|
| Kole | C469F | 2018 | 36 | 0 | 0 | NA | 0 | NA |
| Kole | C469F | 2019 | 39 | 0 | 0 | NA | 0 | NA |
| Kole | C469F | 2020 | 85 | 0 | 0 | NA | 0 | NA |
| Kole | C469F | 2021 | 54 | 0 | 0 | NA | 0 | NA |
| Kole | C469F | 2022 | 72 | 0 | 0 | NA | 0 | NA |
| Kole | R561H | 2016 | 22 | 0 | 0 | NA | 0 | NA |
| Kole | R561H | 2017 | 47 | 0 | 0 | NA | 0 | NA |
| Kole | R561H | 2018 | 36 | 0 | 0 | NA | 0 | NA |
| Kole | R561H | 2019 | 57 | 0 | 0 | NA | 0 | NA |
| Kole | R561H | 2020 | 84 | 0 | 0 | NA | 0 | NA |
| Kole | R561H | 2021 | 42 | 0 | 0 | NA | 0 | NA |
| Kole | R561H | 2022 | 37 | 0 | 0 | NA | 0 | NA |
| Lamwo | P441L | 2016 | 33 | 0 | 0 | NA | 0 | NA |
| Lamwo | P441L | 2017 | 42 | 0 | 0 | NA | 0 | NA |
| Lamwo | P441L | 2018 | 34 | 0 | 0 | NA | 0 | NA |
| Lamwo | P441L | 2019 | 49 | 0 | 0 | NA | 0 | NA |
| Lamwo | P441L | 2020 | 84 | 0 | 0 | NA | 0 | NA |
| Lamwo | P441L | 2021 | 58 | 0 | 0 | NA | 0 | NA |
| Lamwo | P441L | 2022 | 53 | 0 | 0 | NA | 0 | NA |
| Lamwo | C469Y | 2016 | 33 | 3 | 0·09 | 2016 | 7 | 0 |
| Lamwo | C469Y | 2017 | 44 | 3 | 0·07 | 2016 | 7 | 1 |
| Lamwo | C469Y | 2018 | 34 | 8 | 0·24 | 2016 | 7 | 2 |
| Lamwo | C469Y | 2019 | 50 | 5 | 0·1 | 2016 | 7 | 3 |
| Lamwo | C469Y | 2020 | 84 | 28 | 0·33 | 2016 | 7 | 4 |
| Lamwo | C469Y | 2021 | 51 | 16 | 0·31 | 2016 | 7 | 5 |
| Lamwo | C469Y | 2022 | 53 | 11 | 0·21 | 2016 | 7 | 6 |
| Lamwo | A675V | 2016 | 37 | 2 | 0·05 | 2016 | 7 | 0 |
| Lamwo | A675V | 2017 | 44 | 5 | 0·11 | 2016 | 7 | 1 |
| Lamwo | A675V | 2018 | 36 | 1 | 0·03 | 2016 | 7 | 2 |
| Lamwo | A675V | 2019 | 49 | 6 | 0·12 | 2016 | 7 | 3 |
| Lamwo | A675V | 2020 | 84 | 16 | 0·19 | 2016 | 7 | 4 |
| Lamwo | A675V | 2021 | 53 | 12 | 0·23 | 2016 | 7 | 5 |
| Lamwo | A675V | 2022 | 53 | 8 | 0·15 | 2016 | 7 | 6 |
| Lamwo | C469F | 2016 | 31 | 0 | 0 | NA | 0 | NA |
| Lamwo | C469F | 2017 | 43 | 0 | 0 | NA | 0 | NA |
| Lamwo | C469F | 2018 | 31 | 0 | 0 | NA | 0 | NA |
| Lamwo | C469F | 2019 | 31 | 0 | 0 | NA | 0 | NA |
| Lamwo | C469F | 2020 | 84 | 0 | 0 | NA | 0 | NA |
| Lamwo | C469F | 2021 | 51 | 0 | 0 | NA | 0 | NA |
| Lamwo | C469F | 2022 | 46 | 0 | 0 | NA | 0 | NA |
| Lamwo | R561H | 2016 | 32 | 0 | 0 | NA | 0 | NA |
| Lamwo | R561H | 2017 | 44 | 0 | 0 | NA | 0 | NA |
| Lamwo | R561H | 2018 | 31 | 0 | 0 | NA | 0 | NA |
| Lamwo | R561H | 2019 | 50 | 0 | 0 | NA | 0 | NA |
| Lamwo | R561H | 2020 | 84 | 0 | 0 | NA | 0 | NA |
| Lamwo | R561H | 2021 | 42 | 0 | 0 | NA | 0 | NA |
| Lamwo | R561H | 2022 | 37 | 0 | 0 | NA | 0 | NA |
| Mubende | P441L | 2016 | 22 | 0 | 0 | 2020 | 3 | -4 |
| Mubende | P441L | 2017 | 45 | 0 | 0 | 2020 | 3 | -3 |
| Mubende | P441L | 2018 | 38 | 0 | 0 | 2020 | 3 | -2 |
| Mubende | P441L | 2019 | 22 | 0 | 0 | 2020 | 3 | -1 |
| Mubende | P441L | 2020 | 88 | 1 | 0·01 | 2020 | 3 | 0 |
| Mubende | P441L | 2021 | 19 | 1 | 0·05 | 2020 | 3 | 1 |
| Mubende | P441L | 2022 | 52 | 6 | 0·12 | 2020 | 3 | 2 |
| Mubende | C469Y | 2016 | 26 | 0 | 0 | 2017 | 2 | -1 |

| | | | | | | | | |
|---|---|---|---|---|---|---|---|---|
| Mubende | C469Y | 2017 | 46 | 1 | 0·02 | 2017 | 2 | 0 |
| Mubende | C469Y | 2018 | 36 | 0 | 0 | 2017 | 2 | 1 |
| Mubende | C469Y | 2019 | 22 | 0 | 0 | 2017 | 2 | 2 |
| Mubende | C469Y | 2020 | 88 | 3 | 0·03 | 2017 | 2 | 3 |
| Mubende | C469Y | 2021 | 19 | 0 | 0 | 2017 | 2 | 4 |
| Mubende | C469Y | 2022 | 52 | 0 | 0 | 2017 | 2 | 5 |
| Mubende | A675V | 2016 | 28 | 0 | 0 | 2020 | 2 | -4 |
| Mubende | A675V | 2017 | 46 | 0 | 0 | 2020 | 2 | -3 |
| Mubende | A675V | 2018 | 41 | 0 | 0 | 2020 | 2 | -2 |
| Mubende | A675V | 2019 | 24 | 0 | 0 | 2020 | 2 | -1 |
| Mubende | A675V | 2020 | 87 | 6 | 0·07 | 2020 | 2 | 0 |
| Mubende | A675V | 2021 | 19 | 0 | 0 | 2020 | 2 | 1 |
| Mubende | A675V | 2022 | 53 | 6 | 0·11 | 2020 | 2 | 2 |
| Mubende | C469F | 2016 | 26 | 0 | 0 | 2018 | 1 | -2 |
| Mubende | C469F | 2017 | 45 | 0 | 0 | 2018 | 1 | -1 |
| Mubende | C469F | 2018 | 38 | 4 | 0·11 | 2018 | 1 | 0 |
| Mubende | C469F | 2019 | 22 | 0 | 0 | 2018 | 1 | 1 |
| Mubende | C469F | 2020 | 88 | 0 | 0 | 2018 | 1 | 2 |
| Mubende | C469F | 2021 | 19 | 0 | 0 | 2018 | 1 | 3 |
| Mubende | C469F | 2022 | 52 | 0 | 0 | 2018 | 1 | 4 |
| Mubende | R561H | 2016 | 26 | 0 | 0 | NA | 0 | NA |
| Mubende | R561H | 2017 | 46 | 0 | 0 | NA | 0 | NA |
| Mubende | R561H | 2018 | 38 | 0 | 0 | NA | 0 | NA |
| Mubende | R561H | 2019 | 21 | 0 | 0 | NA | 0 | NA |
| Mubende | R561H | 2020 | 88 | 0 | 0 | NA | 0 | NA |
| Mubende | R561H | 2021 | 16 | 0 | 0 | NA | 0 | NA |
| Mubende | R561H | 2022 | 53 | 0 | 0 | NA | 0 | NA |
| Rukiga | P441L | 2016 | 35 | 0 | 0 | 2021 | 1 | -5 |
| Rukiga | P441L | 2017 | 0 | 0 | NA | 2021 | 1 | -4 |
| Rukiga | P441L | 2018 | 0 | 0 | NA | 2021 | 1 | -3 |
| Rukiga | P441L | 2019 | 26 | 0 | 0 | 2021 | 1 | -2 |
| Rukiga | P441L | 2020 | 39 | 0 | 0 | 2021 | 1 | -1 |
| Rukiga | P441L | 2021 | 83 | 1 | 0·01 | 2021 | 1 | 0 |
| Rukiga | P441L | 2022 | 60 | 0 | 0 | 2021 | 1 | 1 |
| Rukiga | C469Y | 2016 | 35 | 0 | 0 | 2022 | 1 | -6 |
| Rukiga | C469Y | 2017 | 0 | 0 | NA | 2022 | 1 | -5 |
| Rukiga | C469Y | 2018 | 0 | 0 | NA | 2022 | 1 | -4 |
| Rukiga | C469Y | 2019 | 26 | 0 | 0 | 2022 | 1 | -3 |
| Rukiga | C469Y | 2020 | 39 | 0 | 0 | 2022 | 1 | -2 |
| Rukiga | C469Y | 2021 | 72 | 0 | 0 | 2022 | 1 | -1 |
| Rukiga | C469Y | 2022 | 46 | 1 | 0·02 | 2022 | 1 | 0 |
| Rukiga | A675V | 2016 | 42 | 0 | 0 | 2019 | 4 | -3 |
| Rukiga | A675V | 2017 | 0 | 0 | NA | 2019 | 4 | -2 |
| Rukiga | A675V | 2018 | 0 | 0 | NA | 2019 | 4 | -1 |
| Rukiga | A675V | 2019 | 21 | 1 | 0·05 | 2019 | 4 | 0 |
| Rukiga | A675V | 2020 | 39 | 1 | 0·03 | 2019 | 4 | 1 |
| Rukiga | A675V | 2021 | 82 | 2 | 0·02 | 2019 | 4 | 2 |
| Rukiga | A675V | 2022 | 59 | 3 | 0·05 | 2019 | 4 | 3 |
| Rukiga | C469F | 2016 | 42 | 8 | 0·19 | 2016 | 4 | 0 |
| Rukiga | C469F | 2017 | 0 | 0 | NA | 2016 | 4 | 1 |
| Rukiga | C469F | 2018 | 0 | 0 | NA | 2016 | 4 | 2 |
| Rukiga | C469F | 2019 | 14 | 0 | 0 | 2016 | 4 | 3 |
| Rukiga | C469F | 2020 | 39 | 11 | 0·28 | 2016 | 4 | 4 |
| Rukiga | C469F | 2021 | 72 | 29 | 0·4 | 2016 | 4 | 5 |
| Rukiga | C469F | 2022 | 60 | 23 | 0·38 | 2016 | 4 | 6 |

| | | | | | | | | |
|---|---|---|---|---|---|---|---|---|
| Rukiga | R561H | 2016 | 41 | 0 | 0 | 2021 | 2 | -5 |
| Rukiga | R561H | 2017 | 0 | 0 | NA | 2021 | 2 | -4 |
| Rukiga | R561H | 2018 | 0 | 0 | NA | 2021 | 2 | -3 |
| Rukiga | R561H | 2019 | 23 | 0 | 0 | 2021 | 2 | -2 |
| Rukiga | R561H | 2020 | 39 | 0 | 0 | 2021 | 2 | -1 |
| Rukiga | R561H | 2021 | 67 | 11 | 0·16 | 2021 | 2 | 0 |
| Rukiga | R561H | 2022 | 53 | 12 | 0·23 | 2021 | 2 | 1 |
| Tororo | P441L | 2016 | 34 | 0 | 0 | NA | 0 | NA |
| Tororo | P441L | 2017 | 48 | 0 | 0 | NA | 0 | NA |
| Tororo | P441L | 2018 | 31 | 0 | 0 | NA | 0 | NA |
| Tororo | P441L | 2019 | 71 | 0 | 0 | NA | 0 | NA |
| Tororo | P441L | 2020 | 40 | 0 | 0 | NA | 0 | NA |
| Tororo | P441L | 2021 | 74 | 0 | 0 | NA | 0 | NA |
| Tororo | P441L | 2022 | 51 | 0 | 0 | NA | 0 | NA |
| Tororo | C469Y | 2016 | 34 | 0 | 0 | 2021 | 2 | -5 |
| Tororo | C469Y | 2017 | 48 | 0 | 0 | 2021 | 2 | -4 |
| Tororo | C469Y | 2018 | 31 | 0 | 0 | 2021 | 2 | -3 |
| Tororo | C469Y | 2019 | 71 | 0 | 0 | 2021 | 2 | -2 |
| Tororo | C469Y | 2020 | 40 | 0 | 0 | 2021 | 2 | -1 |
| Tororo | C469Y | 2021 | 73 | 3 | 0·04 | 2021 | 2 | 0 |
| Tororo | C469Y | 2022 | 51 | 5 | 0·1 | 2021 | 2 | 1 |
| Tororo | A675V | 2016 | 37 | 0 | 0 | 2017 | 3 | -1 |
| Tororo | A675V | 2017 | 48 | 1 | 0·02 | 2017 | 3 | 0 |
| Tororo | A675V | 2018 | 43 | 0 | 0 | 2017 | 3 | 1 |
| Tororo | A675V | 2019 | 73 | 0 | 0 | 2017 | 3 | 2 |
| Tororo | A675V | 2020 | 41 | 0 | 0 | 2017 | 3 | 3 |
| Tororo | A675V | 2021 | 74 | 3 | 0·04 | 2017 | 3 | 4 |
| Tororo | A675V | 2022 | 57 | 9 | 0·16 | 2017 | 3 | 5 |
| Tororo | C469F | 2016 | 35 | 1 | 0·03 | 2016 | 2 | 0 |
| Tororo | C469F | 2017 | 48 | 1 | 0·02 | 2016 | 2 | 1 |
| Tororo | C469F | 2018 | 31 | 0 | 0 | 2016 | 2 | 2 |
| Tororo | C469F | 2019 | 44 | 0 | 0 | 2016 | 2 | 3 |
| Tororo | C469F | 2020 | 40 | 0 | 0 | 2016 | 2 | 4 |
| Tororo | C469F | 2021 | 73 | 0 | 0 | 2016 | 2 | 5 |
| Tororo | C469F | 2022 | 48 | 0 | 0 | 2016 | 2 | 6 |
| Tororo | R561H | 2016 | 34 | 0 | 0 | NA | 0 | NA |
| Tororo | R561H | 2017 | 48 | 0 | 0 | NA | 0 | NA |
| Tororo | R561H | 2018 | 32 | 0 | 0 | NA | 0 | NA |
| Tororo | R561H | 2019 | 70 | 0 | 0 | NA | 0 | NA |
| Tororo | R561H | 2020 | 41 | 0 | 0 | NA | 0 | NA |
| Tororo | R561H | 2021 | 66 | 0 | 0 | NA | 0 | NA |
| Tororo | R561H | 2022 | 39 | 0 | 0 | NA | 0 | NA |

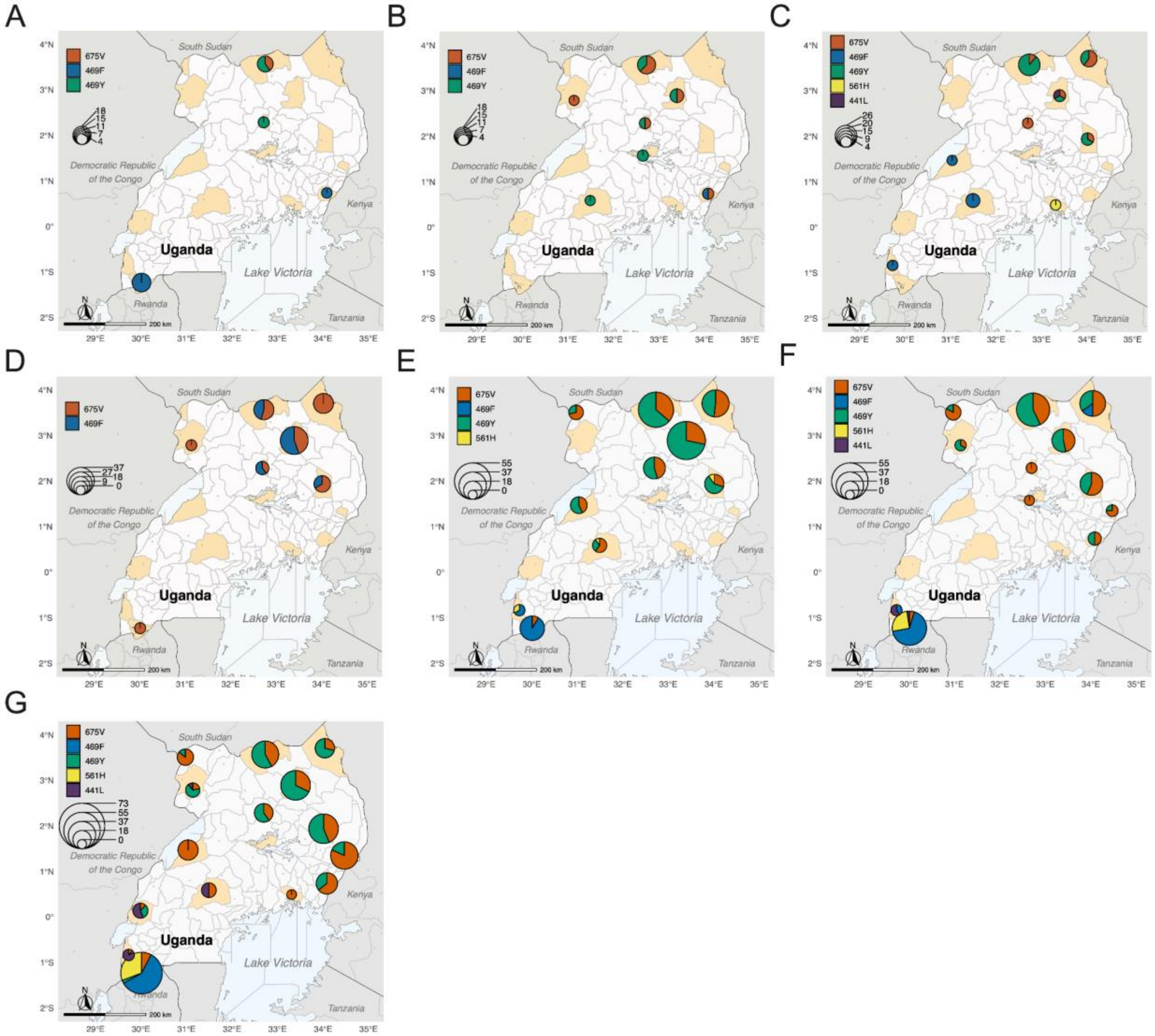


**Figure S2: Evolution of artemisinin partial resistance in Uganda.**
The distribution of mutations and their relative proportion 675V (red), 469F (blue), 469Y (green), 561H (yellow), 441L (purple) across Ugandan sites in 2016 (A), 2017 (B), 2018 (C), 2019 (D), 2020 (E), 2021 (F), and 2022 (G). Study districts are highlighted in orange. The size of the pie charts is scaled by the overall prevalence of all K13 mutations in each district.

### Overview of K13 mutation prevalence data in South-East Asia (SEA)

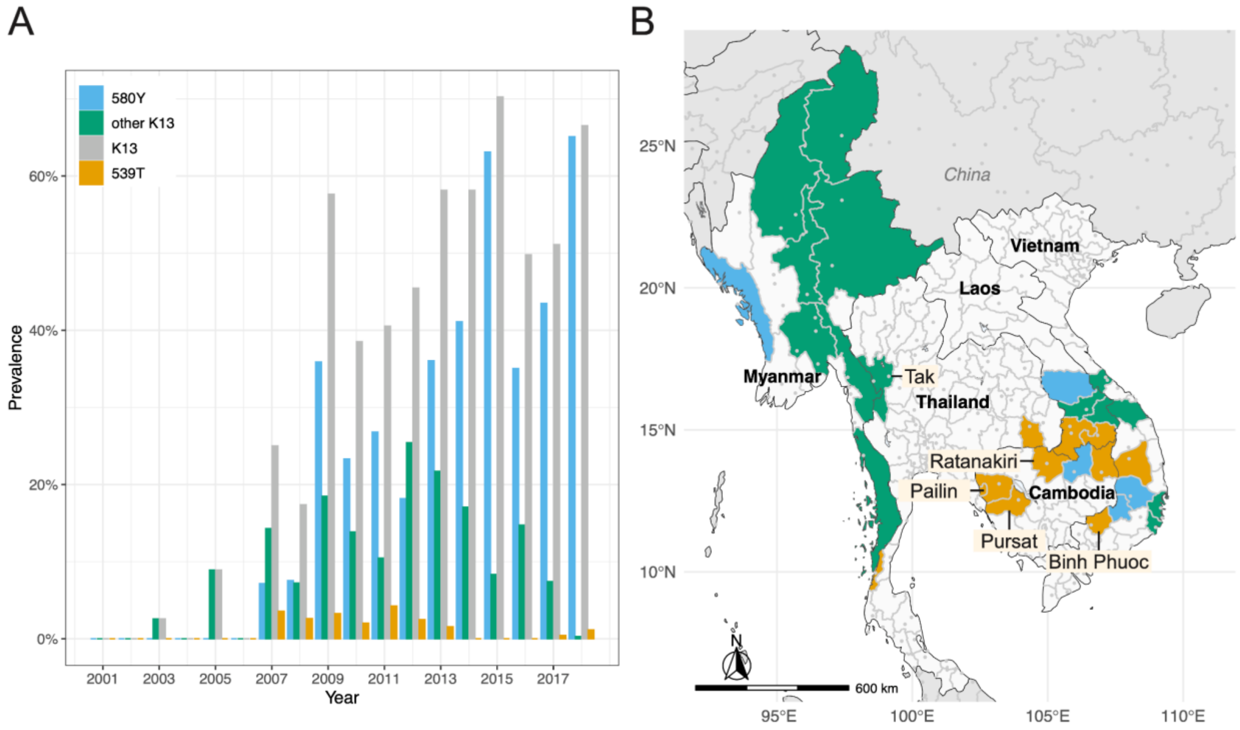


**Figure S3: Evolution of artemisinin partial resistance mutations in SEA.**[2]
(A) Allele frequency of 580Y (blue), 539T (green), other validated K13 propeller domain mutations (orange), and all validated K13 propeller domain mutations (446I, 458Y, 469Y, 476I, 493H, 539T, 543T, 553L, 561H, 574L, 580Y, 622I, 675V) (grey) from 2001 to 2018. (B) The distribution of mutations observed at districts across all time points: 580Y (blue), 580Y, and 539T (orange), other validated K13 propeller domain mutations combined (green) across SEA. Districts for which selection coefficients were estimated are labeled.

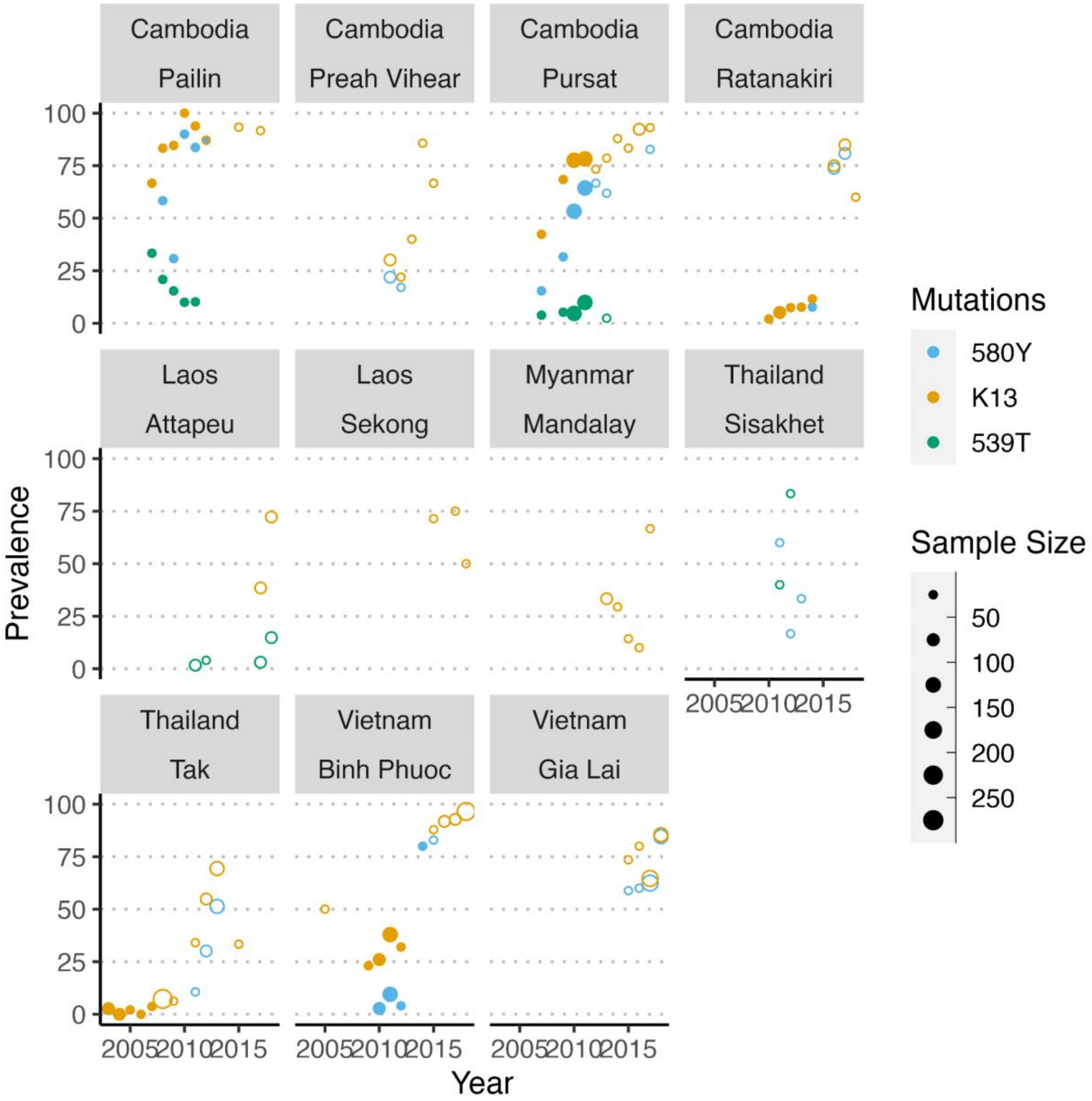


**Figure S4: Overview of prevalence data in SEA observed at least three times at a given site.**
Prevalence of 580Y (blue), 539T (green), and all validated K13 mutations (446I, 458Y, 469Y, 476I, 493H, 539T, 543T, 553L, 561H, 574L, 580Y, 622I, 675V) (orange) in SEA. The filled circles highlight the data used for the selection coefficient estimates. The non-filled circles represent the data that did not pass the three selection criteria (only the first five years of non-zero prevalence with at least three consecutive years, and the first year of non-zero prevalence had to occur before 2011).

### Mixed-effect Bayesian Generalized Linear Model: Estimating selection coefficients in SEA

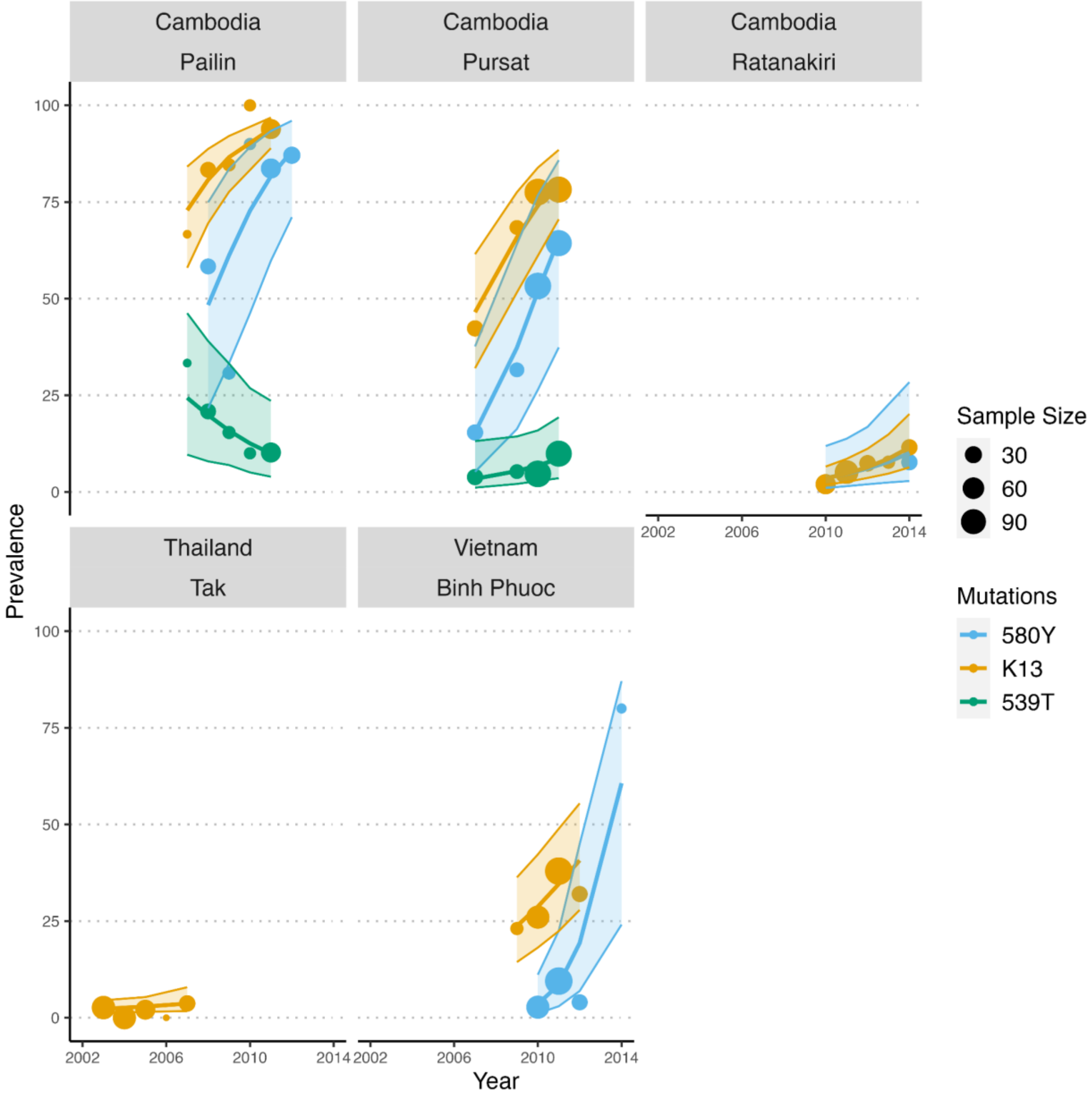


**Figure S5: Selection of kelch13 mutations in SEA.**
Panels show the prevalence of 580Y (blue), 539T (green), and all validated K13 mutations (446I, 458Y, 469Y, 476I, 493H, 539T, 543T, 553L, 561H, 574L, 580Y, 622I, 675V) (orange) over time in each district. Circles indicate the mutation's prevalence and are sized based on the sample size of all genotypes per mutation, district, and year. The lines represent the posterior of a fitted Bayesian Generalized Mixture Model weighted by the inverse variance of the log frequency ratio from each observation. The shaded areas around the lines represent the 95% credible intervals.

### Selection Coefficient estimates in Uganda and SEA

**Table S2: Selection coefficients for Uganda 2016-2022.**

| District | Mutastion | CrI low* | estimate | CrI high ** |
|---|---|---|---|---|
| Agago | 675V | -0·228 | 0·14 | 0·504 |
| Agago | 469Y | -0·188 | 0·244 | 0·663 |
| Agago | K13 | -0·077 | 0·238 | 0·558 |
| Arua | 675V | -0·445 | 0·007 | 0·422 |
| Arua | K13 | -0·038 | 0·339 | 0·714 |
| Hoima | K13 | 0·109 | 0·456 | 0·824 |
| Kaabong | 675V | -0·482 | -0·064 | 0·327 |
| Kaabong | 469Y | -0·273 | 0·205 | 0·589 |
| Kaabong | K13 | -0·242 | 0·106 | 0·439 |
| Kanungu | 469F | -1·12 | 0·125 | 1·363 |
| Kanungu | 441L | 0·195 | 1·235 | 2·431 |
| Kanungu | K13 | 0·283 | 0·644 | 1·057 |
| Kasese | 441L | -0·436 | 0·709 | 1·783 |
| Kasese | K13 | 0·359 | 0·76 | 1·209 |
| Katakwi | 675V | -0·042 | 0·316 | 0·72 |
| Katakwi | 469Y | -0·07 | 0·301 | 0·774 |
| Katakwi | K13 | 0·119 | 0·445 | 0·785 |
| Koboko | 675V | -0·249 | 0·197 | 0·681 |
| Koboko | 469Y | -0·601 | 0·164 | 0·602 |
| Koboko | K13 | -0·168 | 0·265 | 0·681 |
| Kole | 675V | -0·226 | 0·156 | 0·544 |
| Kole | 469Y | -0·231 | 0·198 | 0·576 |
| Kole | K13 | -0·039 | 0·279 | 0·61 |
| Lamwo | 675V | -0·151 | 0·198 | 0·545 |
| Lamwo | 469Y | -0·177 | 0·237 | 0·628 |
| Lamwo | K13 | -0·022 | 0·278 | 0·583 |
| Mubende | 441L | -0·176 | 0·942 | 2·163 |
| Mubende | K13 | -0·01 | 0·333 | 0·68 |
| Rukiga | 675V | -0·362 | 0·117 | 0·578 |
| Rukiga | 469F | -1·06 | 0·182 | 1·426 |
| Rukiga | K13 | 0·109 | 0·415 | 0·717 |
| Tororo | 675V | -0·074 | 0·304 | 0·738 |
| Tororo | K13 | 0·06 | 0·384 | 0·719 |
| Uganda | 469Y | 0·003 | 0·223 | 0·393 |
| Uganda | 675V | -0·009 | 0·155 | 0·322 |
| Uganda | 469F | -0·451 | 0·152 | 0·794 |
| Uganda | 441L | 0·413 | 0·953 | 1·538 |
| Uganda | K13 | 0·243 | 0·379 | 0·527 |

* lower bound of the credible interval (CrI)
** upper bound of the CrI

**Table S3: Minimum and maximum selection coefficient (s) across Uganda per mutation.**

| Mutation | Min selection coefficient | District of min | Max selection coefficient | District of max |
|---|---|---|---|---|
| 675V | -0·064 | Kaabong | 0·316 | Katakwi |
| 469Y | 0·164 | Koboko | 0·301 | Katakwi |
| 469F | 0·125 | Kanungu | 0·182 | Rukiga |
| 441L | 0·709 | Kasese | 1·235 | Kanungu |
| K13 | 0·106 | Kaabong | 0·76 | Kasese |

**Table S4: Selection coefficients for SEA 2003-2018.**

| District | Mutastion | CrI low* | estimate | CrI high ** |
|---|---|---|---|---|
| Binh Phuoc | 580Y | -0·331 | 0·912 | 2·317 |
| Pailin | 580Y | -0·787 | 0·523 | 1·806 |
| Pursat | 580Y | -0·712 | 0·584 | 1·884 |
| Ratanakiri | 580Y | -1·057 | 0·286 | 1·545 |
| Pailin | 539T | -1·968 | -0·263 | 1·39 |
| Pursat | 539T | -1·413 | 0·235 | 1·913 |
| Binh Phuoc | K13 | -0·2 | 0·269 | 0·723 |
| Pailin | K13 | -0·03 | 0·434 | 0·902 |
| Pursat | K13 | -0·052 | 0·392 | 0·85 |
| Ratanakiri | K13 | -0·128 | 0·309 | 0·804 |
| Tak | K13 | -0·397 | 0·136 | 0·56 |
| Southeast Asia | 580Y | -0·092 | 0·574 | 1·201 |
| Southeast Asia | 539T | -0·852 | -0·005 | 0·814 |
| Southeast Asia | K13 | 0·089 | 0·308 | 0·536 |

* lower bound of the CrI
** upper bound of the CrI

**Table S5: Minimum and maximum selection coefficient (s) across SEA per mutation.**

| Mutation | Min selection coefficient | District of min | Max selection coefficient | District of max |
|---|---|---|---|---|
| 580Y | 0·286 | Ratanakiri | 0·912 | Binh Phuoc |
| 539T | -0·263 | Pailin | 0·235 | Pursat |
| K13 | 0·136 | Tak | 0·434 | Pailin |

### Selection Coefficient estimates in Uganda (2016-2021)

Using the mixed-effects Bayesian Generalized Linear model, we also estimated selection coefficients for each kelch13 mutation and an overall selection of total kelch mutants per site (the total number of infections with any validated kelch mutations per site and year) for Uganda from 2016-2021, excluding the last year of sampling (2022).

**Table S6: Selection Coefficients for Uganda 2016-2021.**

| District | Mutastion | CrI low* | estimate | CrI high ** |
|---|---|---|---|---|
| Agago | 675V | -0·146 | 0·263 | 0·691 |
| Agago | 469Y | 0·034 | 0·467 | 0·931 |
| Agago | K13 | 0·149 | 0·424 | 0·718 |
| Arua | 675V | -0·571 | 0·01 | 0·426 |
| Arua | K13 | -0·156 | 0·237 | 0·531 |
| Hoima | K13 | 0·15 | 0·405 | 0·979 |
| Kaabong | 675V | -0·28 | 0·213 | 0·637 |
| Kaabong | 469Y | -0·137 | 0·357 | 0·781 |
| Kaabong | K13 | 0·097 | 0·406 | 0·701 |
| Kanungu | 469F | -1·189 | 0·121 | 1·405 |
| Kanungu | 441L | -0·066 | 1·044 | 2·204 |
| Kanungu | K13 | 0·002 | 0·324 | 0·674 |
| Kasese | 441L | -0·113 | 1·039 | 2·138 |
| Kasese | K13 | 0·136 | 0·396 | 0·943 |
| Katakwi | 675V | -0·114 | 0·288 | 0·76 |
| Katakwi | 469Y | -0·117 | 0·349 | 0·78 |
| Katakwi | K13 | 0·101 | 0·389 | 0·692 |
| Koboko | 675V | -0·273 | 0·259 | 0·849 |
| Koboko | 469Y | -0·386 | 0·317 | 0·804 |
| Koboko | K13 | -0·031 | 0·372 | 0·708 |
| Kole | 675V | -0·197 | 0·221 | 0·673 |
| Kole | 469Y | -0·184 | 0·297 | 0·733 |
| Kole | K13 | 0·07 | 0·341 | 0·63 |
| Lamwo | 675V | -0·112 | 0·284 | 0·7 |
| Lamwo | 469Y | -0·033 | 0·393 | 0·811 |
| Lamwo | K13 | 0·171 | 0·415 | 0·678 |
| Mubende | 441L | -0·013 | 1·044 | 2·317 |
| Mubende | K13 | -0·06 | 0·303 | 0·564 |
| Rukiga | 675V | -0·639 | 0·044 | 0·467 |
| Rukiga | 469F | -1·07 | 0·206 | 1·467 |
| Rukiga | K13 | 0·132 | 0·402 | 0·665 |
| Tororo | 675V | -0·373 | 0·104 | 0·507 |
| Tororo | K13 | -0·086 | 0·248 | 0·508 |
| Uganda | 469Y | 0·128 | 0·361 | 0·552 |
| Uganda | 675V | -0·029 | 0·188 | 0·358 |
| Uganda | 469F | -0·524 | 0·157 | 0·752 |
| Uganda | 441L | 0·441 | 1·042 | 1·638 |
| Uganda | K13 | 0·246 | 0·36 | 0·483 |

* lower bound of the CrI
** upper bound of the CrI

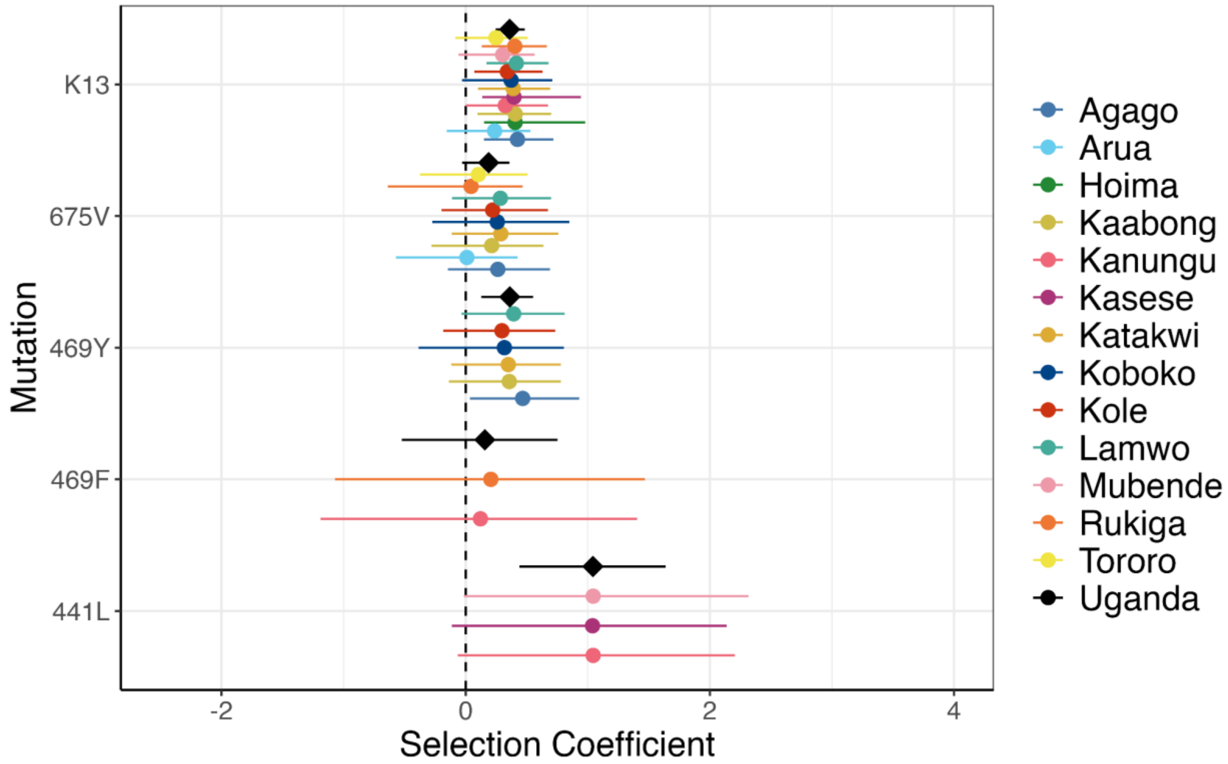


**Figure S6: Estimated selection coefficients per year in Uganda 2016-2021.**
Per year point estimates for individual sites (colored circles) or all combined sites (black diamonds) are shown with lines representing the 95% CrI for the indicated mutations in Uganda (A) and SEA (B), including the combined mutations (K13 combined).

### Literature Review

#### Table S7: Comparison of selection coefficients obtained from a literature review

| Study | Selection coefficient | Protein | Mutation | Front-Line Drug | Location | Timeframe |
|---|---|---|---|---|---|---|
| Anderson and Roper (2005)[3] | 0·150 | triple-mutant DHFR | 51I/59R, 108N | Pyrimethamine | South Africa | 1995-1999 |
| Anderson and Roper (2005)[3] | 0·228 | double-mutant DHFR | 51I/108N; 59R/108N | Pyrimethamine | South Africa | 1995-1999 |
| Anderson and Roper (2005)[3] | 0·390 | double mutant DHPS | 437G/540E | Pyrimethamine | South Africa | 1995-1999 |
| Nair et al. (2003)[4] | 0·660 | DHFR | 108N | Pyrimethamine | Thailand/ Myanmar | 1975-1981 |
| Nwakanma et al. (2014)[5] | 0·300 | CRT | 76T | Chloroquine | Gambia | 1984-2008 |
| Nwakanma et al. (2014)[5] | 0·260 | MDR | 86Y | Chloroquine | Gambia | 1984-2008 |
| Nwakanma et al. (2014)[5] | 0·220 | DFR | 51I, 59R, 108N | Sulfadoxine-Pyrimethamine | Gambia | 1984-2008 |
| Nwakanma et al. (2014)[5] | 0·220 | DHPS | 437G | Sulfadoxine-Pyrimethamine | Gambia | 1984-2008 |
| Nsanzabana et al. (2010)[6] | 0·156 | CRT | C72S | Chloroquine | Papua New Guinea | 1991-2002 |
| Nsanzabana et al. (2010)[6] | 0·245 | CRT | K76T | Chloroquine | Papua New Guinea | 1991-2002 |
| Nsanzabana et al. (2010)[6] | 0·368 | CRT | H97Q | Chloroquine | Papua New Guinea | 1991-2002 |
| Nsanzabana et al. (2010)[6] | 0·152 | CRT | I356L | Chloroquine | Papua New Guinea | 1991-2002 |
| Nsanzabana et al. (2010)[6] | -0·296 | CRT | I356T | Chloroquine | Papua New Guinea | 1991-2002 |
| Nsanzabana et al. (2010)[6] | 0·129 | CRT | A220S | Chloroquine | Papua New Guinea | 1991-2002 |
| Nsanzabana et al. (2010)[6] | 0·102 | CRT | N326D | Chloroquine | Papua New Guinea | 1991-2002 |
| Nsanzabana et al. (2010)[6] | 0·062 | MDR1 | N86Y | Chloroquine | Papua New Guinea | 1991-2002 |
| Nsanzabana et al. (2010)[6] | -0·076 | MDR1 | Y184F | Chloroquine | Papua New Guinea | 1991-2002 |
| Nsanzabana et al. (2010)[6] | 0·196 | MDR1 | N1042D | Chloroquine | Papua New Guinea | 1991-2002 |
| Nsanzabana et al. (2010)[6] | 0·135 | DHFR | C59R | Sulfadoxine-Pyrimethamine | Papua New Guinea | 1991-2002 |
| Nsanzabana et al. (2010)[6] | 0·117 | DHFR | S108N | Sulfadoxine-Pyrimethamine | Papua New Guinea | 1991-2002 |
| Okell et al. (2018)[7] | 0·153 | MDR1 | 184F | AL [1] | Africa | 1990-2018 |
| Okell et al. (2018)[7] | 0·033 | MDR1 | 184F | AL/AS-AQ [2] | Africa | 1990-2018 |
| Okell et al. (2018)[7] | 0·069 | MDR1 | 184F | AS-AQ [3] | Africa | 1990-2018 |
| Okell et al. (2018)[7] | -0·249 | MDR1 | 1246Y | AL [1] | Africa | 1990-2018 |
| Okell et al. (2018)[7] | -0·105 | MDR1 | 1246Y | AL/AS-AQ [2] | Africa | 1990-2018 |
| Okell et al. (2018)[7] | 0·063 | MDR1 | 1246Y | AS-AQ [3] | Africa | 1990-2018 |
| Okell et al. (2018)[7] | 0·165 | MDR1 | 86Y | AL [1] | Africa | 1990-2018 |
| Okell et al. (2018)[7] | -0·210 | MDR1 | 86Y | AL/AS-AQ [2] | Africa | 1990-2018 |
| Okell et al. (2018)[7] | -0·204 | MDR1 | 86Y | AS-AQ [3] | Africa | 1990-2018 |
| Anderson et al. (2016)[8] | 0·960 | K13 | 580Y | Artemisinin | Thailand/ Myanmar | 2001-2014 |
| Anderson et al. (2016)[8] | 0·480 | K13 | combined *** | Artemisinin | Thailand/ Myanmar | 2001-2014 |
| Meier-Scherling et al. (2024) | 0·222 | K13 | 469Y | Artemisinin | Uganda | 2016-2021 |
| Meier-Scherling et al. (2024) | 0·152 | K13 | 675V | Artemisinin | Uganda | 2016-2021 |
| Meier-Scherling et al. (2024) | 0·153 | K13 | 469F | Artemisinin | Uganda | 2016-2021 |
| Meier-Scherling et al. (2024) | 0·968 | K13 | 441L | Artemisinin | Uganda | 2016-2021 |
| Meier-Scherling et al. (2024) | 0·383 | K13 | K13 **** | Artemisinin | Uganda | 2016-2021 |
| Meier-Scherling et al. (2024) | 0·574 | K13 | 580Y | Artemisinin | Southeast Asia | 1991-2015 |
| Meier-Scherling et al. (2024) | -0·005 | K13 | 539T | Artemisinin | Southeast Asia | 1991-2015 |
| Meier-Scherling et al. (2024) | 0·308 | K13 | K13 ***** | Artemisinin | Southeast Asia | 1991-2015 |

[1] Artemether–lumefantrine
[2] Artesunate–amodiaquine
[3] Artemether–lumefantrine or artesunate-amodiaquine
*** A481V, A675V, C580Y, D281V, E252Q, F446I, F614L, G449A, G533A, G538V, K438N, K479I, M476I, N458Y, N537I, P441L, P527H, P527L, P553L, P553P, P574L, P667Q, P667R, P667T, R239Q, R515K, R528G, R561H, R575K, S485N, Y511H
**** 469Y, 675V, 469F, *441L
***** 580Y, 446I, 458Y, 476I, 493H, 539T, 543T, 553L, 553S, 561H

### Forecasting selection using mixed-effect Bayesian Generalized Linear Model

We estimated selection coefficients based on the first three, four, and five years of mutation prevalence data after the first non-zero mutation occurred in each district using the same mixed-effects framework. Subsequently, we forecast the prevalence of each resistance mutation from the first three, four, and five years up until 2023 using the posterior prediction assuming constant selection (Figure S8).

Based on the method described in Figure S7, we forecasted the selection of all validated mutations (Figure S8A) and 580Y (Figure S8B) for each district in SEA. To assess the forecasting accuracy, the correlation, bias, and mean average error (MAE) between all data, depicted as points in Figure S8A-B, and the forecasted selection, represented by dashed lines in Figure S8A-B, was calculated.

| Ratanakiri, Cambodia | | | | | | |
|---|---|---|---|---|---|---|
| **Year** | **Sample size** | **K13** | **Prevalence** | **Forecasting** | | |
| | | | | *3 years* | *4 years* | *5 years* |
| 2010 | 50 | 1 | 2.00 | | | |
| 2011 | 78 | 4 | 5.13 | | | |
| 2012 | 27 | 2 | 7.41 | | | |
| 2013 | 13 | 1 | 7.69 | 11.26 | | |
| 2014 | 26 | 3 | 11.54 | 16.5 | 11.19 | |
| 2015 | | | | 23.87 | 14.65 | 13.27 |
| 2016 | 80 | 60 | 75.00 | 33.25 | 18.5 | 16.62 |
| 2017 | 73 | 62 | 84.93 | 43.7 | 23.21 | 20.38 |
| 2018 | 5 | 3 | 60.00 | 55.41 | 28.89 | 24.32 |
| 2019 | | | | 66.31 | 35.81 | 29.58 |
| 2020 | | | | 75.67 | 41.9 | 34.31 |
| 2021 | | | | 83.22 | 49.85 | 40.55 |
| 2022 | | | | 88.6 | 57.31 | 46.62 |
| 2023 | | | | 92.5 | 64.2 | 53.04 |

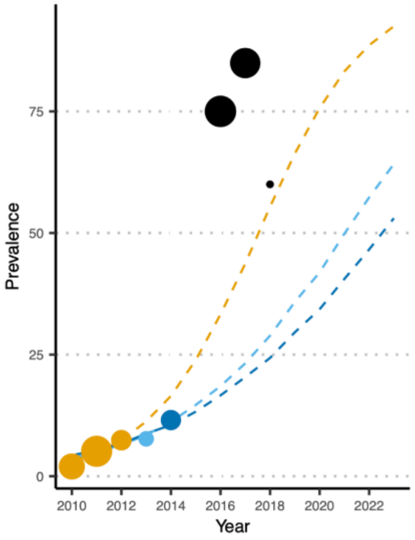


**Figure S7. Overview of forecasted selection of artemisinin resistance mutations in SEA.**
Example of forecasting the selection of all combined K13 mutations until the year 2023 in Ratanakiri, Cambodia, based on the first three (orange), four (light blue), and five (dark blue) years (2010-2014) of non-zero prevalence (orange). (A) First, the Bayesian mixed-effect linear models was fit to data from 2010-2014. (B) Then, the selection was forecasted until 2023 (grey) based on the first five years of non-zero prevalence (orange).

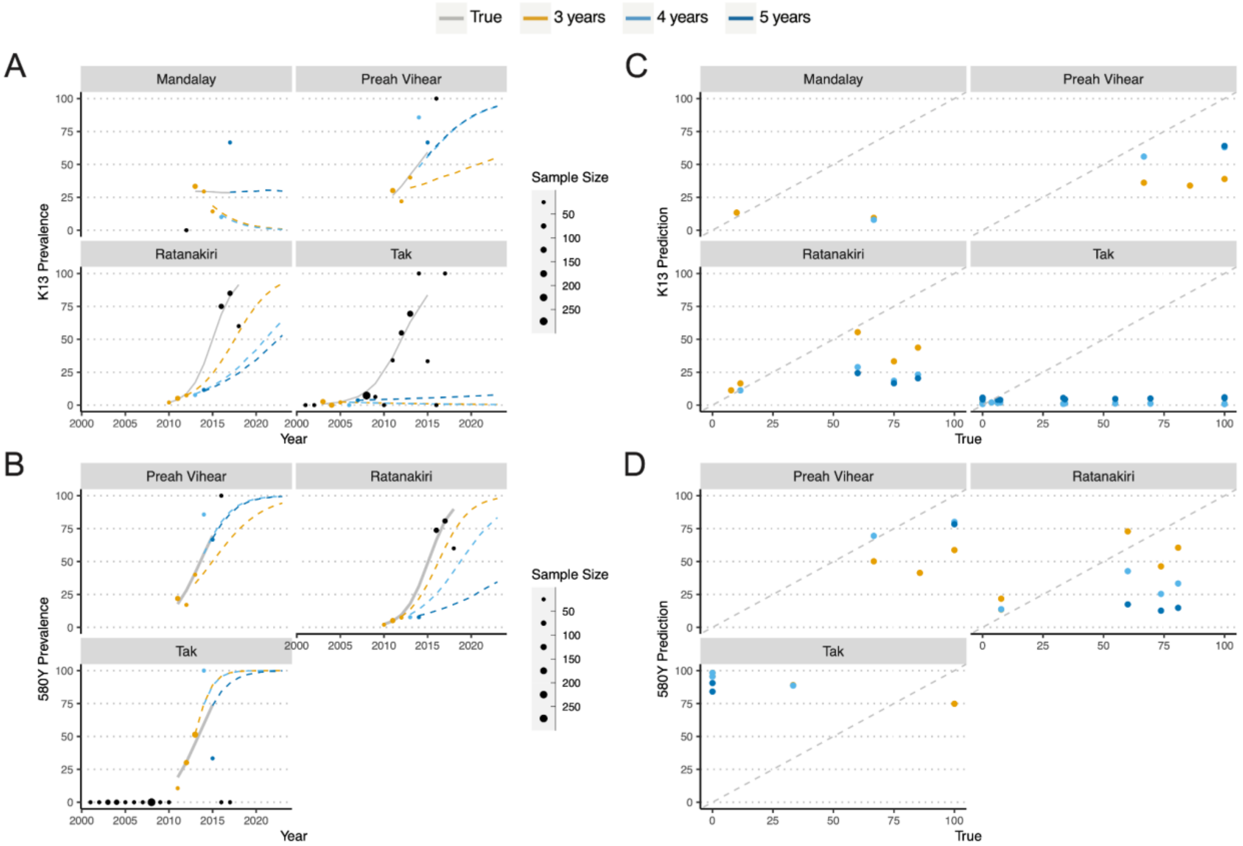


**Figure S8. Forecasting selection of partial artemisinin resistance mutations in SEA.**
The selection of 580Y (A) and 13 validated K13 propeller domain mutations (B) was forecasted based on the first three (orange), four (light blue), and five (dark blue) years in SEA. (C) The correlation between all data (true) and the forecasted selection (predicted) for all validated K13 propeller domain mutations was r = 0·47, 0·45, 0·48 for the three (orange), four (light blue), and five (dark blue) year predictions, respectively. (D) The correlation between all data (true) and the forecasted selection (predicted) of 580Y was r = -0·08, -0·29, -0·57 for the three (orange), four (light blue), and five (dark blue) year predictions, respectively.

For each forecasted selection of either 580Y or all K13 mutations based on the first 3, 4, or 5 years of non-zero prevalence, the mean average error (MAE), bias, and correlation estimates were calculated. The MAE is calculated as the mean(abs(true-prediction). The bias is calculated as the mean(true - prediction).

**Table S8: Analysis of forecasting based on the first 3, 4, or 5 years of non-zero prevalence in SEA**

| Locus | # year used for forecasting | MAE | Bias | Correlation |
|---|---|---|---|---|
| 580Y | 3 | 38·12 | -8·95 | -0·08 |
| 580Y | 4 | 43·44 | -13·88 | -0·29 |
| 580Y | 5 | 60·98 | 2·78 | -0·57 |
| K13 | 3 | 31·87 | 30·37 | 0·47 |
| K13 | 4 | 36·41 | 36·15 | 0·45 |
| K13 | 5 | 40·78 | 39·34 | 0·48 |
